# Turbulent dynamics and in silico perturbation-based measures are disrupted across biological stages of Alzheimer’s disease

**DOI:** 10.64898/2026.10.12.26365318

**Authors:** Noelia Martínez-Molina, David Aquilué-Llorens, Lars Rass, Guillermo Montaña-Valverde, Gustavo Patow, Anira Escrichs, Yonatan Sanz-Perl, Morten L. Kringelbach, Satu Palva, Fernando de Maestú, Gustavo Deco, the Alzheimer’s Disease Neuroimaging Initiative

**Affiliations:** Computational Neuroscience Group, Center for Brain and Cognition, Department of Information and Communication Technologies, Universitat Pompeu Fabra, Barcelona, Catalonia, Spain; Digital Health Unit, Eurecat Technology Center, Barcelona, Spain; Neuroscience Center, HiLIFE-Helsinki Institute of Life Science, University of Helsinki, Helsinki, Finland; ViRVIG, Universitat de Girona, Girona, Catalonia, Spain; Center for Music in the Brain, Department of Clinical Medicine, Aarhus University, Aarhus, Denmark; Centre for Eudaimonia and Human Flourishing, University of Oxford, Oxford, OX1 2JD, UK; Centre for Cognitive Neuroimaging, School of Psychology and Neuroscience, University of Glasgow, Glasgow; Centre of Cognitive and Computational Neuroscience, Universidad Complutense de Madrid (UCM), Madrid, Spain; Institució Catalana de la Recerca i Estudis Avançats (ICREA), Barcelona, Catalonia, Spain

**Keywords:** computational modelling, turbulent dynamics, machine learning, Alzheimer’s Disease

## Abstract

According to the AT(N) framework, Alzheimer’s disease (AD) can be biologically characterised by amyloid deposition, tau pathology, and neurodegeneration, yet how these processes relate to whole-brain dynamics remains unclear. Here, we investigated these relationships and the link between whole-brain dynamics and cognitive dysfunction using multimodal neuroimaging from 145 ADNI3 participants. We combined multiscale measures of turbulent dynamics with responses to in silico perturbations obtained from whole-brain models. AD progression was characterised by a scale-dependent reorganisation of turbulent dynamics where information flow showed the strongest contribution to the dynamics-related variance in cognitive performance. Large-scale turbulence was associated with all three AT(N) components. Model-based information capability and susceptibility measures progressively decreased across AD stages and were associated with amyloid burden and regional tau deposition. Model-free features discriminated comparisons involving AD dementia, whereas adding model-based measures produced modest, metric-dependent changes in classification performance. Exploratory analyses showed spatial alignment between large-scale turbulence alterations and AD-risk gene-expression gradients, with *APOE* among the strongest associations in both disease-stage contrasts examined. Lastly, we found a progressive decoupling between turbulent dynamics and the E:I balance across AD stages. Together, these findings link model-based and model-free measures of whole-brain dynamics to proteinopathy, neurodegeneration, and cognitive impairment, supporting their complementary value for characterising AD progression.

## INTRODUCTION

Alzheimer’s disease (AD) is the leading cause of dementia, which affects more than 55 million people worldwide (Alzheimer’s Association 2024). The diagnostic and staging criteria for AD have evolved substantially, guided largely by the International Working Group (Dubois et al. 2024) and the Alzheimer’s Association (Jack et al. 2024). Both frameworks now incorporate a biological definition of the disease. Biologically, AD is characterised by the pathological accumulation of amyloid-*β* (A*β*), including soluble oligomers and extracellular plaques associated with synaptic dysfunction (Busche and Konnerth 2016), as well as intracellular neurofibrillary tangles composed of tau that disrupt neuronal structure and function (Busche et al. 2019). Importantly, A*β* and tau accumulation precede the development of neurodegeneration and cognitive symptoms (Therriault et al. 2022, 2024). Current Core 1 biomarkers include A*β* positron emission tomography (PET), cerebrospinal fluid measures of A*β*42, and phosphorylated tau proteins (Jack et al. 2024). Blood-based biomarkers such as phosphorylated tau 217 provide scalable and minimally invasive indices of cerebral amyloid pathology, including during preclinical stages when tau PET abnormalities may not yet be detectable (Janelidze et al. 2021, Palmqvist et al. 2017). Blood samples are also routinely used for APOE *ε*4 genotyping and the assessment of other AD-associated genetic variants (Sepulcre et al. 2018). However, current biological definitions do not incorporate direct measures of complex brain dynamics, despite the fundamental role of these dynamics in supporting brain function and cognition.

Resting-state fMRI (rsfMRI) is a noninvasive technique that measures BOLD activity and places little cognitive demand on the participants which makes it especially suitable for AD patients. The human brain operates as a complex dynamical system in which activity emerges from interactions among distributed regions across multiple spatial and temporal scales. A central feature of this organisation is metastability (Deco and Kringelbach 2016, Tognoli and Kelso 2014), whereby the brain continuously transitions between states of integration and segregation, enabling flexible coordination among neural populations. Whole-brain computational modelling has provided a mechanistic basis for this view, demonstrating that rsfMRI activity is best captured near regimes of maximal metastability, where local and global interactions are balanced and the system can flexibly explore a rich repertoire of configurations (Deco et al. 2017).

Crucially, metastability has been reported to be decreased in AD (Córdova-Palomera et al. 2017, Demirtaş et al. 2017). Recently, we have extended the concept of metastability to characterise multiscale organisation of brain dynamics inspired by turbulence (Deco and Kringelbach 2020). Our turbulent dynamics framework captures the variability of local synchronisation across spatial scales using the local Kuramoto order parameter. In AD, evidence from animal models has found neuronal hyperactivity near amyloid plaques that could be explained by inhibitory dysfunction (Busche et al. 2008, Busche and Konnerth 2015, Palop and Mucke 2016) while hypersynchronisation of neuronal oscillations predicts the progression from MCI to AD (Maestú et al. 2021, Pusil et al. 2019). Additionally, evidence from MEG studies further indicates that the AD continuum is characterised by disruptions in functional E:I balance that shift the brain towards more supercritical dynamical regimes (Javed et al. 2025). Taken together, these findings suggest that pathological processes in AD could manifest in scale-dependent alterations in brain dynamics. In this context, a multiscale turbulent framework provides a unique opportunity to characterise how dynamical changes unfold across spatial scales along the AD continuum.

Furthermore, whole-brain computational modelling studies have begun to provide mechanistic insights into how the underlying pathology alters large-scale brain dynamics. Notably, our group demonstrated that AD-related changes can be captured by shifts in the local bifurcation parameter of whole-brain Hopf models, providing a mechanistic explanation for alterations in functional connectivity and brain dynamics across the AD continuum (Demirtaş et al. 2017). These findings support the view that AD is associated with a displacement of the brain from its optimal dynamical regime. In parallel, we also showed that different dementia etiologies exhibit different perturbational trajectories as a function of the in silico stimulation protocol (Sanz Perl et al. 2023). Collectively, these findings highlight the potential of generative modelling to capture AD-related alterations in brain dynamics.

In the present study, we test the hypothesis that AD is characterised by a progressive alteration of whole-brain dynamical regimes that can be associated with the AT(N) biomarkers and cognitive impairment with a framework combining model-free and model-based measures. Using multimodal neuroimaging from the ADNI3 cohort, we combine multiscale measures of turbulent brain dynamics with perturbational measures obtained from whole-brain Hopf models to analyse changes across the AD continuum. For whole-brain modelling, we used the generative effective connectivity (GEC) matrix. The GEC is an extension of the classic concept of effective connectivity (Friston et al. 2003), but generative in the sense that the GEC is created using a linearized Hopf whole-brain model of empirical rsfMRI data, where the strengths of existing anatomical connectivity are adapted iteratively until best fit, thus providing a mechanistic explanation of the rsfMRI data. We then assess whether these dynamical measures are associated with cognitive impairment as well as amyloid burden, tau deposition, and hippocampal volume. We also explored the spatial alignment between alterations in turbulent dynamics and AD-risk gene expression patterns. Finally, we examined whether the relationship between large-scale turbulent dynamics and E:I balance, with a regional transcriptomic proxy, was altered across the AD continuum. By integrating multiscale descriptions of turbulent dynamics and in silico perturbational measures with the AT(N) framework and cognitive impairment, the present work establishes a foundation for characterising the systems-level mechanisms of disease progression, which may inform future neuroimaging markers for patient management and monitoring in real-world settings.

## RESULTS

Figure 1 summarises our methodological pipeline. Our neuroimaging sample from the ADNI3 database included 145 individuals, with 51 healthy controls A*β* negative (HC-) and 37 A*β* positive (HC+), 31 mild cognitive impairment patients A*β* positive (MCI+) and 26 Alzheimer’s disease A*β* positive patients (AD+). Table 1 shows demographic and clinical characteristics for this cohort. There were no significant differences in age or sex across groups but AD+ patients had fewer education years than both healthy-control groups. Primary statistical analyses controlled for sex, age and years of education as a proxy for cognitive reserve. Figure S1 displays mean regional accumulation of A*β* in the brain (in Centiloid, CL, units) stratified by diagnostic group.

**Table 1.** Demographic and clinical characteristics of the ADNI3 cohort. (N = 145 after harmonisation and exclusion of motion outliers). Values are mean *±*SD for continuous variables and counts for categorical variables. Group comparisons were performed using Kruskal–Wallis tests for continuous variables and Pearson’s chi-square test for sex. Significant continuous-variable omnibus tests were followed by Dunn pairwise tests with Holm correction. Effect sizes for significant continuous-variable omnibus tests are reported as epsilon squared (*ε*^2^). ^a^*p*_Holm_ *<* 0.05 vs HC^*−*^; ^b^*p*_Holm_ *<* 0.05 vs HC^+^; ^c^*p*_Holm_ *<* 0.05 vs MCI^+^; ^d^*p*_Holm_ *<* 0.05 vs AD^+^. *Abbreviations:* MMSE, Mini-Mental State Examination; MOCA, Montreal Cognitive Assessment; ADAS-Cog 13, Alzheimer’s Disease Assessment Scale–Cognitive Subscale (13-item version).

| Variable | HC<sup>−</sup><br>(n = 51) | HC<sup>+</sup><br>(n = 37) | MCI<sup>+</sup><br>(n = 31) | AD<sup>+</sup><br>(n = 26) | Test | p-value | Effect size |
| --- | --- | --- | --- | --- | --- | --- | --- |
| Age (years) | 72.47 ± 5.27 | 74.76 ± 5.75 | 73.45 ± 6.33 | 75.85 ± 7.56 | KW | 0.071 | – |
| Sex (M/F) | 27 / 24 | 16 / 21 | 13 / 18 | 16 / 10 | $\chi^2$ | 0.386 | – |
| Education (years) | 16.84 ± 2.28<sup>d</sup> | 16.86 ± 2.30<sup>d</sup> | 16.03 ± 2.40 | 14.65 ± 2.48<sup>a,b</sup> | KW | 0.002 | 0.086 |
| MMSE Memory | 9.65 ± 0.63<sup>c,d</sup> | 9.68 ± 0.53<sup>c,d</sup> | 8.81 ± 1.01<sup>a,b,d</sup> | 6.96 ± 1.80<sup>a,b,c</sup> | KW | < 0.001 | 0.437 |
| MMSE Overall | 29.06 ± 1.14<sup>c,d</sup> | 28.89 ± 1.17<sup>c,d</sup> | 26.87 ± 1.86<sup>a,b,d</sup> | 22.27 ± 2.46<sup>a,b,c</sup> | KW | < 0.001 | 0.603 |
| MOCA | 26.20 ± 2.25<sup>b,c,d</sup> | 24.67 ± 2.54<sup>a,c,d</sup> | 21.65 ± 3.09<sup>a,b,d</sup> | 15.23 ± 4.44<sup>a,b,c</sup> | KW | < 0.001 | 0.581 |
| ADAS-Cog 13 | 9.49 ± 5.23<sup>c,d</sup> | 11.10 ± 4.63<sup>c,d</sup> | 22.18 ± 8.92<sup>a,b,d</sup> | 37.61 ± 11.75<sup>a,b,c</sup> | KW | < 0.001 | 0.608 |
| Centiloid (Aβ) | 2.02 ± 10.27<sup>b,c,d</sup> | 79.84 ± 41.80<sup>a</sup> | 100.27 ± 44.56<sup>a</sup> | 108.33 ± 42.12<sup>a</sup> | KW | < 0.001 | 0.700 |

**Figure 1.**
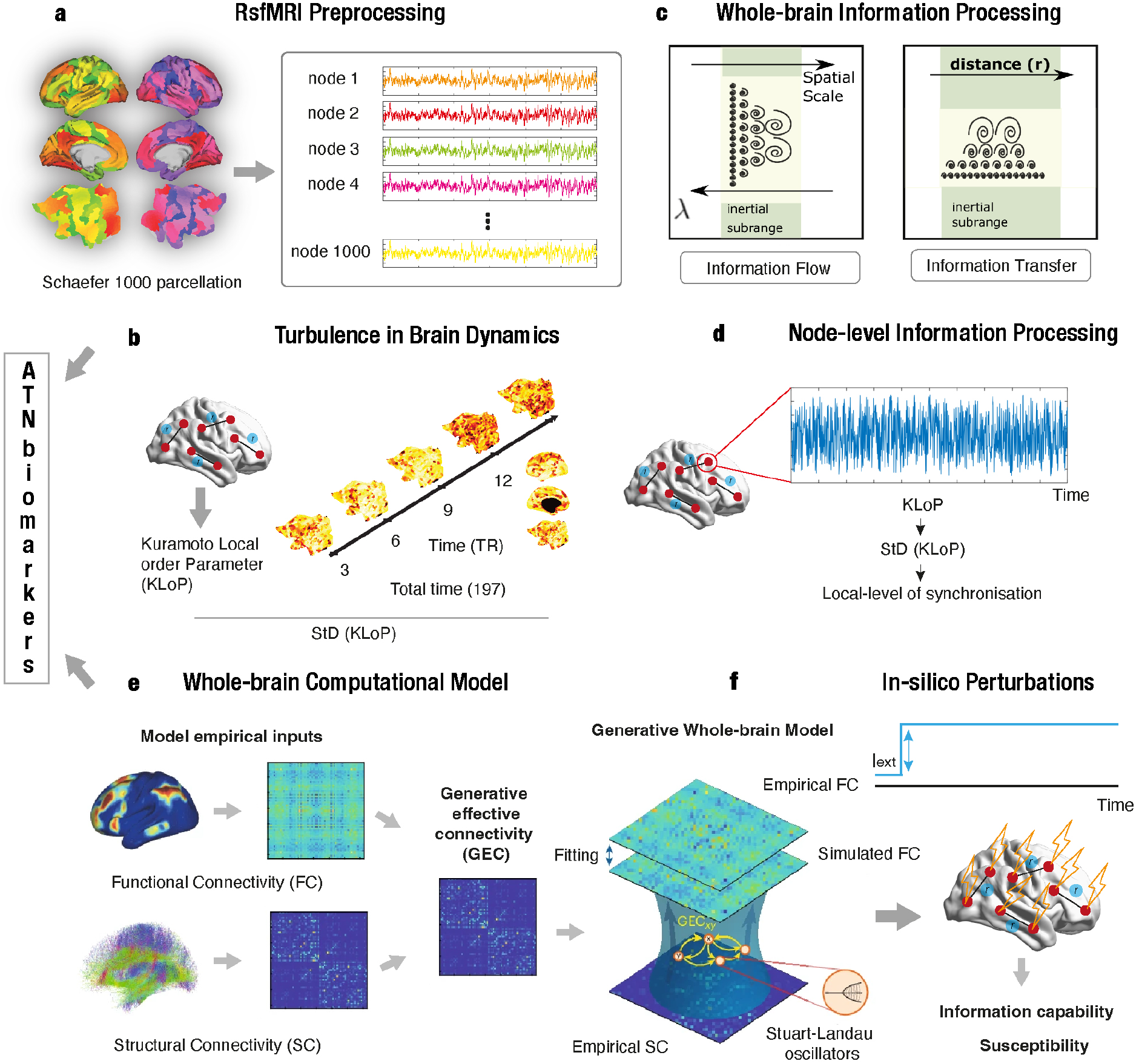
Overview of the turbulent-dynamics and whole-brain modelling framework. (a) Preprocessed rsfMRI time series were extracted using the Schaefer 1,000-node Yeo 7 resting-state networks parcellation and Hilbert-transformed into phase information. (b) Turbulence was quantified from the spatiotemporal variability of the local Kuramoto order parameter across spatial scales (*λ* = 0.01–0.27). (c) Whole-brain information processing was characterised using information flow across successive spatial scales and information transfer between spatial scales. (d) Node-level information processing was quantified from the temporal variability of the local Kuramoto order parameter. (e) Whole-brain Hopf models were constructed from structural and functional connectivity. Generative effective connectivity (GEC) was iteratively fitted so that the coupled Stuart–Landau oscillators reproduced statistical observables from the empirical rsfMRI data (Deco et al. 2017). We implemented both group-level and subject-level whole-brain Hopf models. (f) External stimuli were simulated through *in silico* perturbations, and the model response was quantified using information capability and susceptibility.

### AD alterations in turbulent dynamics are scale-dependent

We first examined how the spatiotemporal patterns of turbulent dynamics varied across AD stages. In addition, we included measures of information flow, information cascade, and information transfer. After data harmonisation, adjustment for age, sex, and education, and FDR correction (see Methods), we found a scale-dependent dissociation in turbulent dynamics between HC^−^and AD^+^.

At lower values of *λ* (*λ* = 0.06, 0.03, and 0.01; Fig. 2a), i.e. large spatial scales, turbulent dynamics were significantly reduced in AD^+^ compared with HC^−^ (*p*_FDR_ = 0.0284, 0.0032, and 0.00245, respectively). In contrast, turbulent dynamics were significantly elevated in AD^+^ at *λ* = 0.27 (Fig. 2b; *p*_FDR_ = 0.00345). When comparing MCI^+^ and AD^+^, turbulent dynamics were significantly reduced in AD^+^ only at *λ* = 0.01 (Fig. 2a; *p*_FDR_ = 0.0128; *g* = 0.82); no other spatial scale survived FDR correction. Comparisons between HC^−^ and HC^+^, HC^+^ and MCI^+^, and HC^−^ and MCI^+^ did not reach statistical significance. All pairwise results are summarised in Table S1.

**Figure 2.**
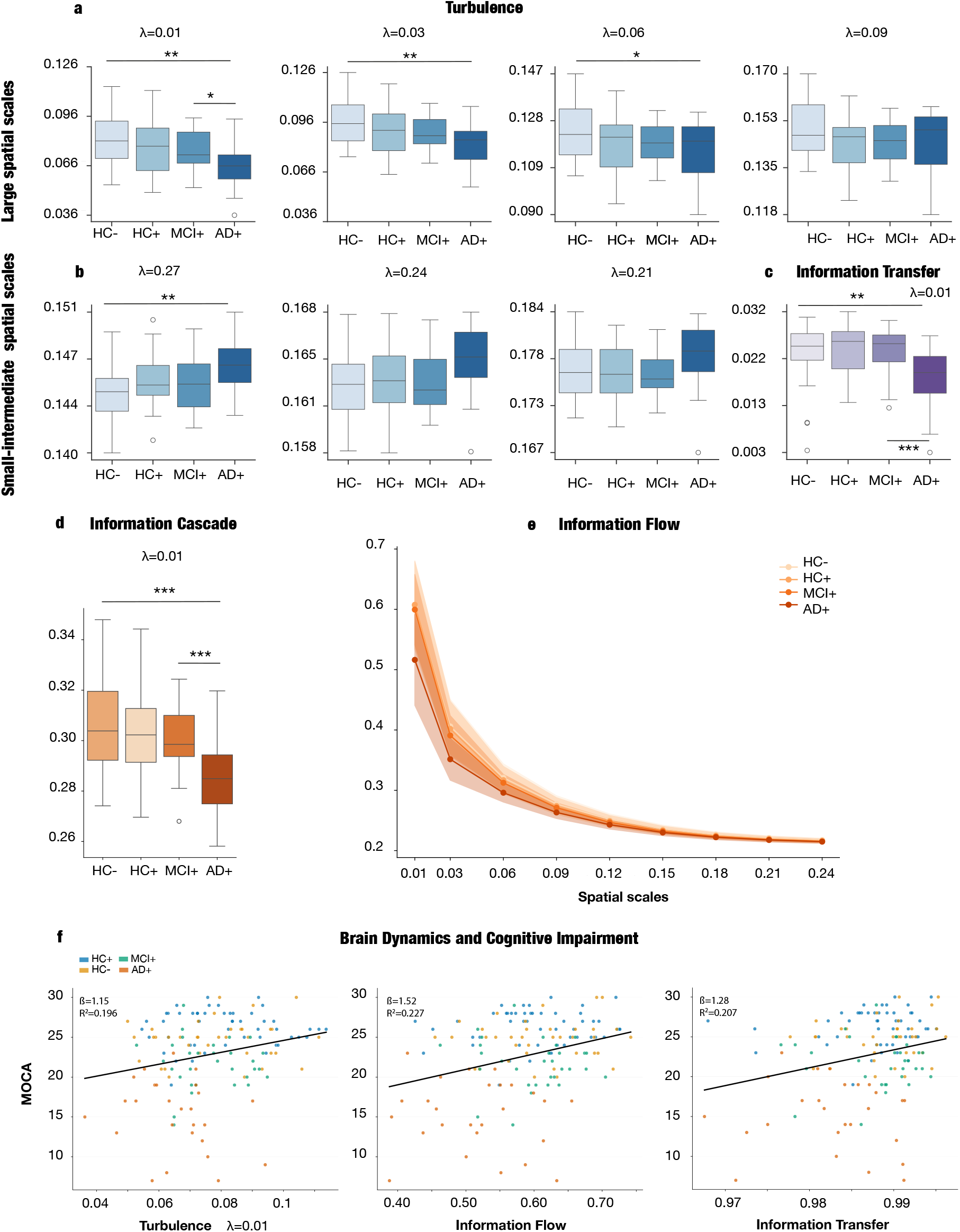
Multiscale alterations of turbulent dynamics and information processing across the AD continuum. (a) Group differences in turbulence at the larger spatial scales *λ* = 0.01, 0.03, 0.06, and 0.09. (b) Turbulence at the smaller spatial scales *λ* = 0.27, 0.24, and 0.21. Boxplots in (a–d) display medians and interquartile ranges; asterisks indicate FDR-corrected pairwise differences (^∗^*p <* 0.05, ^*∗∗*^*p <* 0.01, ^*∗∗∗*^*p <* 0.001). (c) Information transfer at *λ* = 0.01, showing lower values in AD^+^ than in in HC and MCI+. (d) Information cascade at *λ* = 0.01, showing a reduction along the AD continuum. (e) Information flow across spatial scales (*λ* = 0.01–0.24). Lines show group means and shaded regions show variability across participants. (f) Associations of MoCA score with turbulence, information flow, and information transfer at *λ* = 0.01. Points represent individual participants and colours indicate diagnostic group. Solid lines show fitted linear associations; regression coefficients (*β*) and full-model *R*^2^ values are reported within each panel.

Information transfer (Fig. 2c), information cascade (Fig. 2d), and information flow (Fig. 2e) showed systematic differences. AD^+^ participants showed significantly lower information flow than HC^−^ participants at *λ* = 0.21–0.01, and lower information transfer at *λ* = 0.15–0.01. Relative to MCI^+^, AD^+^ participants also showed significantly lower information flow and information transfer at *λ* = 0.09–0.01. Information cascade was significantly lower in AD^+^ than in both MCI^+^ and HC^−^ participants. In addition, MCI^+^ participants showed significantly lower information flow than HC^−^ participants at *λ* = 0.24–0.12. No other comparisons survived FDR correction (Table S2).

### Information flow explains the largest dynamics-related variance in cognitive performance

We next examined whether large-scale brain dynamics were associated with cognitive performance, using MoCA as a measure of global cognition. Separate multiple linear regression models evaluated turbulence, information flow and information transfer at *λ* = 0.01, while controlling for age, sex and education. Information flow showed the strongest association, with each standard-deviation increase corresponding to a 1.52-point increase in MoCA score (*β* = 1.52, *p*_FDR_ = 0.0016), and explained an additional 8.0% of variance beyond the covariates. Significant positive associations were also observed for information transfer (*β* = 1.28, *p*_FDR_ = 0.0085; Δ*R*^2^ = 0.060) and turbulence (*β* = 1.15, *p*_FDR_ = 0.0031; Δ*R*^2^ = 0.049) (Fig. 2f). To account for the shared variance among these highly correlated dynamical measures, we additionally performed a descriptive LMG relative-importance decomposition. Information flow accounted for the largest proportion of the joint dynamics-related explained variance (50.7%), followed by information transfer (28.1%) and turbulence (21.2%).

### Node-level metastability reveals network-level disruptions across the AD continuum

We next examined node-level metastability to characterise fine-grained alterations in information-processing dynamics. Based on the whole-brain analysis, node-wise comparisons were performed at *λ* = 0.01, which showed significant differences between HC^−^ and AD^+^ and between MCI^+^ and AD^+^. After adjustment for age, sex and education, AD^+^ participants showed lower node-level metastability than HC^−^ participants at all 1,000 nodes, all of which survived FDR correction across Schaefer nodes. The MCI^+^ vs. AD^+^ contrast showed a similarly widespread but more selective pattern, with 917 of 1,000 nodes remaining significant after FDR correction. Figure 3a–b displays the spatial distribution of the group differences after adjusting for covariates.

**Figure 3.**
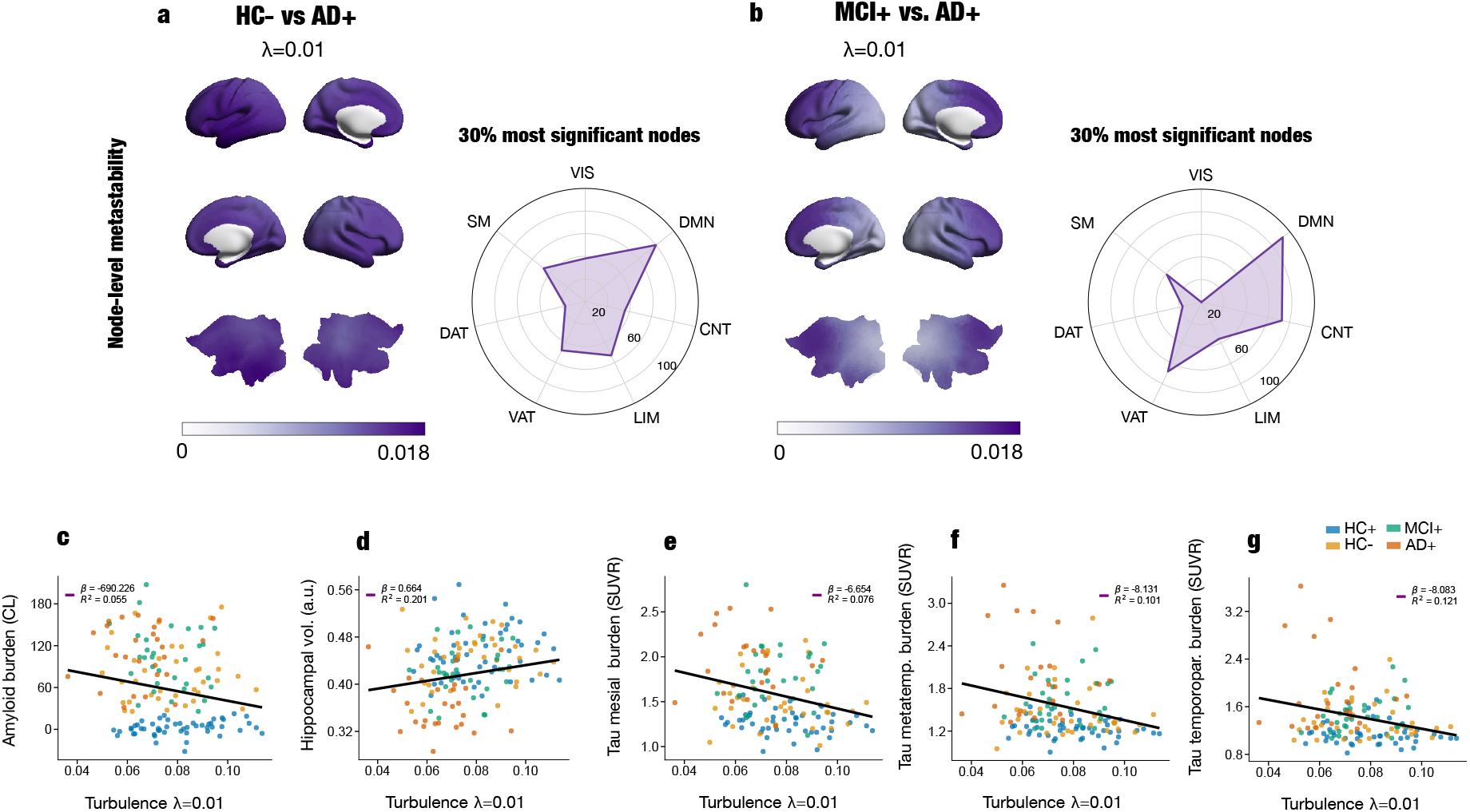
Node-level metastability differences and associations between large-scale turbulence and AT(N) biomarkers. (a,b) Node-level metastability differences at the largest spatial scale (*λ* = 0.01) for the HC^*−*^ vs. AD^+^ (a) and MCI^+^ vs. AD^+^ (b) contrasts. Brain renderings display group mean differences after adjustment for covariates. Radar plots summarise the distribution of nodes within the 30% most statistically significant regions across the Yeo 7 resting-state networks (VIS, SM, DAT, VAT, LIM, CNT, and DMN). (c–g) Associations between turbulence at *λ* = 0.01 and AT(N) biomarkers. Lower turbulence was associated with greater cortical amyloid burden in Centiloid units (c), smaller bilateral hippocampal volume (d), and greater tau burden in mesial temporal (e), metatemporal (f), and temporoparietal (g) regions, expressed as standardised uptake value ratios (SUVR). Points represent individual participants and colours indicate diagnostic group. Solid lines show fitted linear associations; regression coefficients (*β*) and full-model *R*^2^ values are reported within each panel.

To characterise the distribution of the strongest effects across large-scale networks, we selected nodes at or below the 30th percentile of the FDR-adjusted permutation *p*-value distribution and assigned them to the Yeo 7 resting-state networks. This resulted in 320 selected nodes for HC^−^ vs. AD^+^ and 325 nodes for MCI^+^ vs. AD^+^. The DMN contained the largest absolute number of selected nodes in both contrasts, comprising 80 nodes (25.0%) in HC^−^ vs. AD^+^ and 92 nodes (28.3%) in MCI^+^ vs. AD^+^.

### Model-based measures distinguish between AD stages

To characterise the dynamical regime underlying AD progression, we quantified the response of group-level whole-brain Hopf models to external perturbations using measures of information capability and susceptibility with the Schaefer 1000-node parcellation. For each AD stage, perturbation responses were estimated across repeated simulation trials, and statistical differences were assessed using permutation testing on the resulting trial distributions.

Both information capability and susceptibility showed robust differences across AD stages (Fig. 4a–b). Information capability differed significantly across all pairwise stage comparisons after FDR correction, including early transitions between HC^−^ and HC^+^ (*p*_FDR_ = 0.001), intermediate transitions such as HC^+^ vs. MCI^+^ (*p*_FDR_ = 0.04), and the late transition between MCI^+^ and AD^+^ (*p*_FDR_ = 0.001). Comparisons between HC^−^ and both MCI^+^ and AD^+^ were also statistically significant (all *p*_FDR_ = 0.001).

**Figure 4.**
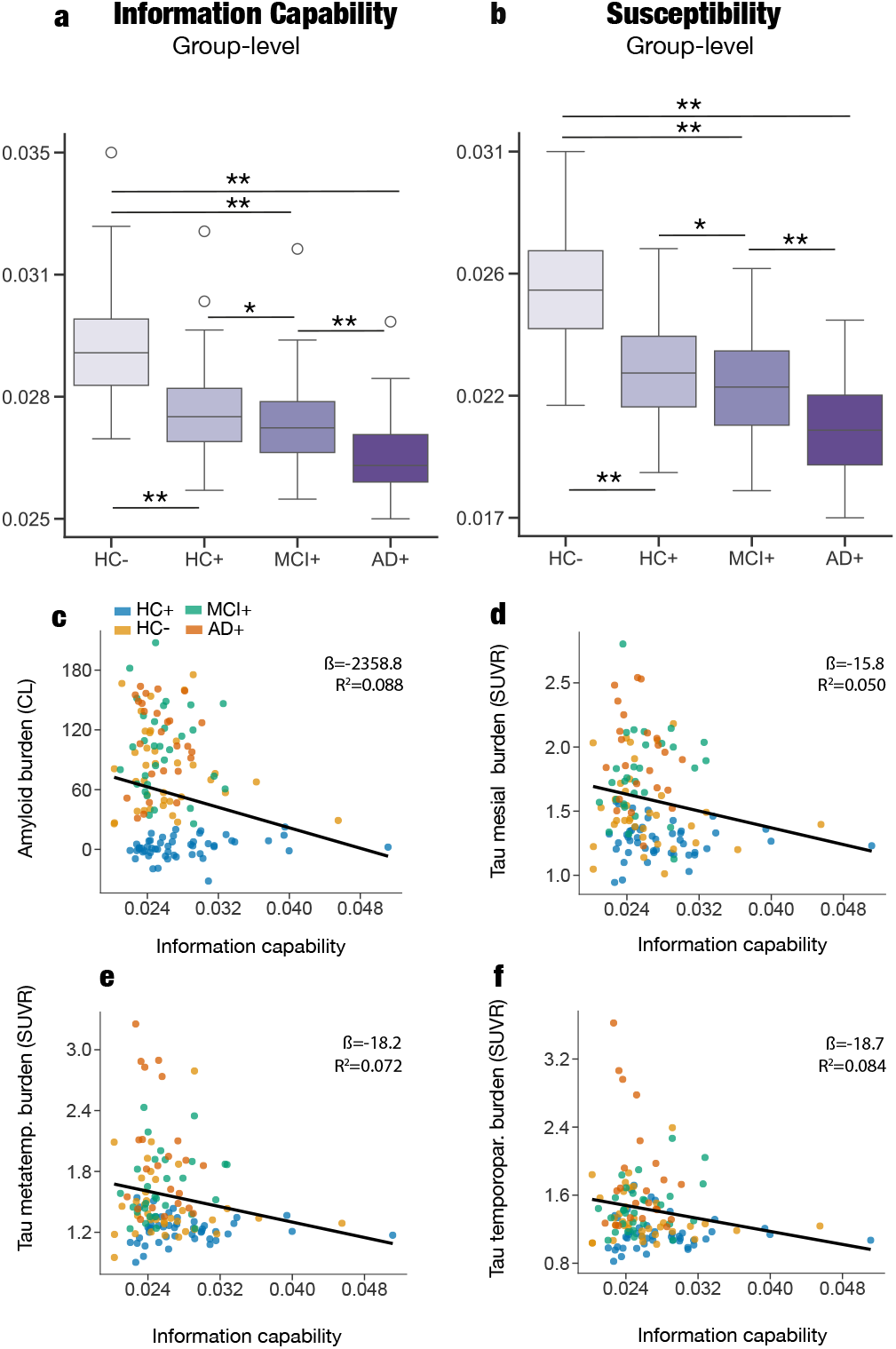
Whole-brain model-based measures track AD staging and are linked to proteinopathy. (a,b) Group-level information capability (a) and susceptibility (b) across HC^*−*^, HC^+^, MCI^+^, and AD^+^, showing progressive reductions along the AD continuum. Boxplots display medians and interquartile ranges; asterisks indicate FDR-corrected pairwise differences (^∗^*p <* 0.05, ^*∗∗*^*p <* 0.01). (c–f) Associations between participant-level information capability and cortical amyloid burden in Centiloid units (c), mesial temporal tau burden (d), metatemporal tau burden (e), and temporoparietal tau burden (f). Tau burden is expressed as standardised uptake value ratio (SUVR). Points represent individual participants and colours indicate diagnostic group. Solid lines show fitted linear associations; regression coefficients (*β*) and full-model *R*^2^ values are reported within each panel.

Similarly, susceptibility exhibited significant differences across all pairwise contrasts (all *p*_FDR_ ≤ 0.030), indicating a consistent alteration in the system’s responsiveness to perturbations throughout disease progression.

Of note, the perturbation measures described above were derived from group-level Hopf models and evaluated across simulation trials; therefore, they do not directly quantify inter-individual variability. To determine whether these model-based measures also varied across individuals along the AD continuum, we estimated information capability and susceptibility using subject-specific Hopf models and compared the resulting participant-level measures using permutation tests adjusted for age, sex, and years of education (Fig. S2). The largest differences were observed between HC^−^ and AD^+^ participants, with higher information capability and susceptibility in HC^−^. Although the direction of the effects was consistent with the group-level simulations, these effects did not reach significance after FDR correction (*p*_FDR_ = 0.165 and 0.184, respectively). The weaker statistical evidence at the participant level may reflect greater variability in subject-specific model estimates and the variance accounted for by the demographic covariates.

We additionally examined whether the subject-level perturbation measures were associated with cognitive performance, as assessed using MoCA scores. Neither information capability nor susceptibility was significantly associated with cognition after controlling for age, sex, and education (information capability: *β* = 0.438, *p*_FDR_ = 0.168; susceptibility: *β* = 0.440, *p*_FDR_ = 0.168).

### Turbulence and model-free measures of brain dynamics are associated with AT(N) biomarkers

To determine whether individual differences in whole-brain dynamics were associated with AT(N) biomarkers, we performed multiple linear regression analyses examining cortical amyloid burden, regional tau deposition, and bilateral hippocampal grey matter volume. Information capability, susceptibility, and turbulence at the largest spatial scale (*λ* = 0.01) were evaluated in separate models controlling for age, sex, and years of education. Statistical significance was assessed using FDR correction across the five prespecified AT(N) outcomes separately for each variable. Associations with amyloid and tau burden are shown in Fig. 3c–g and Fig. 4c–f, with the corresponding susceptibility associations provided in Fig. S2.

All three measures were significantly associated with cortical amyloid burden. Higher information capability and susceptibility were associated with lower amyloid deposition (information capability: *β* = −2358.83, *p*_FDR_ = −0.0112; susceptibility: *β* = −1280.29, *p*_FDR_ = −0.0139). Similarly, lower turbulence was associated with greater amyloid burden (*β* = −649.70, *p*_FDR_ = 0.0392).

Associations with neurodegeneration were examined using bilateral hippocampal grey matter volume. Information capability and susceptibility showed positive associations with hippocampal volume, but neither survived FDR correction (information capability: *β* = 1.44, *p*_FDR_ = 0.1145; susceptibility: *β* = 0.81, *p*_FDR_ = 0.1075). Finally, all three measures were consistently and negatively associated with regional tau deposition across areas corresponding to successive Braak stages. Higher information capability was associated with lower tau burden in mesial temporal (*β* = −15.79, *p*_FDR_ = 0.0172), metatemporal (*β* = −18.16, *p*_FDR_ = 0.0222), and temporoparietal regions (*β* = −18.67, *p*_FDR_ = 0.0327). Susceptibility showed comparable associations in mesial temporal (*β* = −8.62, *p*_FDR_ = 0.0197), metatemporal (*β* = −9.99, *p*_FDR_ = 0.0252), and temporoparietal regions (*β* = −10.29, *p*_FDR_ = 0.0362). Turbulence was similarly associated with lower tau deposition in mesial temporal (*β* = −6.56, *p*_FDR_ = 0.0160), metatemporal (*β* = −7.97, *p*_FDR_ = 0.0221), and temporoparietal regions (*β* = −7.98, *p*_FDR_ = 0.0310).

### Machine learning classification of AD stages based on whole-brain dynamics

We trained logistic regression classifiers across four pairwise classification scenarios: HC^−^ vs. HC^+^, HC^−^ vs. MCI^+^, HC^−^ vs. AD^+^, and MCI^+^ vs. AD^+^. Each scenario was evaluated using two feature specifications:

(1) empirical turbulent-dynamics measures and (2) empirical turbulent-dynamics measures combined with the subject-level information capability and susceptibility estimates (Fig. 5).

**Figure 5.**
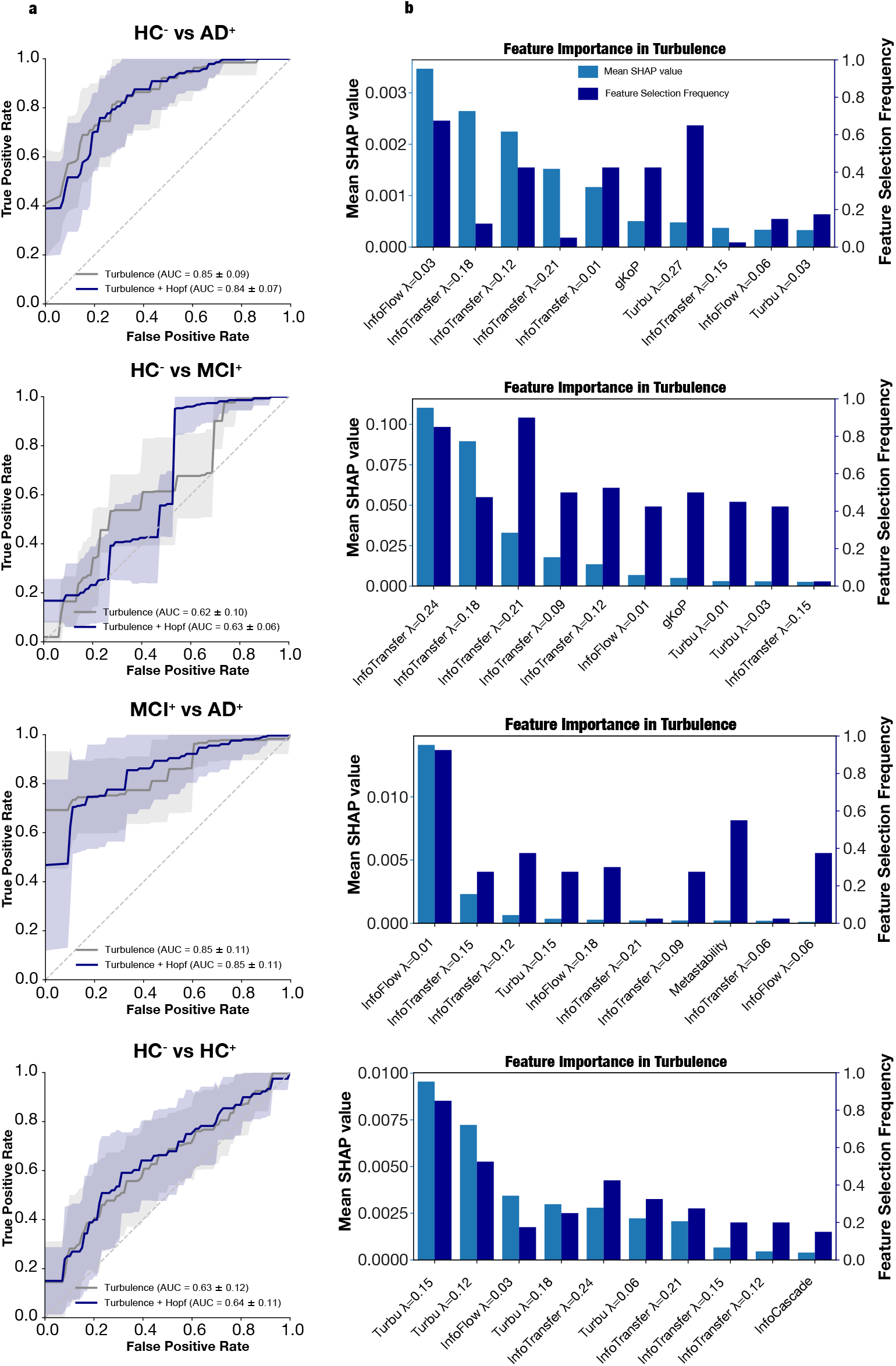
Machine-learning classification and feature importance across disease stages. Results are shown, from top to bottom, for HC^*−*^ vs. AD^+^, HC^*−*^ vs. MCI^+^, MCI^+^ vs. AD^+^, and HC^*−*^ vs. HC^+^. (a) Receiver operating characteristic (ROC) curves for logistic-regression classifiers trained using empirical turbulent-dynamics features alone (grey) or together with information capability and susceptibility from the Hopf models (blue). Solid lines show the mean ROC curves across 40 repeated train-test partitions, shaded regions show the corresponding standard deviations, and mean ± SD area under the curve (AUC) values are reported in each panel. (b) Feature-importance profiles for the empirical turbulent-dynamics classifiers. Light-blue bars report the mean absolute SHapley Additive exPlanations (SHAP) value in held-out test samples, and dark-blue bars report feature-selection frequency across the 40 repetitions. Features are ordered by mean absolute SHAP value within each comparison; *λ* denotes spatial scale.

We performed 40 repeated random 75%/25% train/test partitions to reduce sensitivity to a single data split. Within each outer training set, mRMR feature selection and leave-one-out cross-validation (LOOCV) were used to optimise the number of retained features and the logistic-regression hyperparameters. In the combined specification, information capability and susceptibility were always retained, whereas mRMR was applied to the eligible empirical dynamics features. The selected features and optimal hyperparameters were then used to train a final model on the complete training set, which was evaluated on the corresponding held-out test set. Performance metrics were aggregated across the 40 repetitions and are reported as mean ± SD. Summary results for all contrasts and feature specifications are provided in Table 2.

**Table 2.** Classification performance for turbulent dynamics features and combined turbulent dynamics and perturbation-based features across disease-stage contrasts. Values are reported as mean ± standard deviation across 40 resampling iterations. Performance metrics are expressed as percentages.

| Feature Set | N Selected Features | ROC-AUC (Test) | Balanced Accuracy (Test) | Sensitivity (Test) | Specificity (Test) |
| --- | --- | --- | --- | --- | --- |
| **HC ($A\beta^-$) vs. AD ($A\beta^+$)** |  |  |  |  |  |
| Turbulence | $8.55 \pm 2.96$ | $84.95 \pm 9.05$ | $73.39 \pm 6.31$ | $64.66 \pm 16.39$ | $82.13 \pm 11.31$ |
| Turbulence + Perturbation | $7.83 \pm 2.67$ | $83.98 \pm 7.25$ | $75.02 \pm 8.46$ | $72.56 \pm 15.57$ | $77.48 \pm 12.38$ |
| **HC ($A\beta^-$) vs. MCI ($A\beta^+$)** |  |  |  |  |  |
| Turbulence | $9.60 \pm 1.73$ | $61.86 \pm 9.59$ | $58.41 \pm 12.86$ | $65.38 \pm 17.55$ | $51.44 \pm 12.98$ |
| Turbulence + Perturbation | $7.72 \pm 2.04$ | $63.18 \pm 6.23$ | $55.26 \pm 7.46$ | $52.57 \pm 7.62$ | $57.94 \pm 10.30$ |
| **MCI ($A\beta^+$) vs. AD ($A\beta^+$)** |  |  |  |  |  |
| Turbulence | $9.78 \pm 2.93$ | $85.18 \pm 10.73$ | $66.37 \pm 12.59$ | $74.03 \pm 18.28$ | $58.72 \pm 20.19$ |
| Turbulence + Perturbation | $8.43 \pm 3.22$ | $84.58 \pm 10.57$ | $68.22 \pm 11.15$ | $59.93 \pm 28.93$ | $76.50 \pm 22.13$ |
| **HC ($A\beta^-$) vs. HC ($A\beta^+$)** |  |  |  |  |  |
| Turbulence | $10.45 \pm 3.11$ | $63.07 \pm 12.25$ | $59.45 \pm 10.15$ | $47.72 \pm 22.56$ | $71.19 \pm 21.34$ |
| Turbulence + Perturbation | $10.35 \pm 3.89$ | $64.34 \pm 11.05$ | $59.58 \pm 12.74$ | $51.67 \pm 21.29$ | $67.49 \pm 11.92$ |

Using empirical turbulent-dynamics features alone, the strongest discrimination was observed for comparisons involving AD^+^. HC^−^ vs. AD^+^ classification achieved an AUC of 84.95 ± 9.05% and a balanced accuracy of 73.39 ±6.31%. MCI^+^ vs. AD^+^ produced a similarly high AUC of 85.18 ±10.73%, although its balanced accuracy was lower and more variable (66.37 ± 12.59%). Performance was more modest for HC^−^ vs. MCI^+^ (AUC = 61.86 ± 9.59%; balanced accuracy = 58.41 ±12.86%) and for the preclinical HC^−^ vs. HC^+^ contrast (AUC = 63.07 ± 12.25%; balanced accuracy = 59.45 ± 10.15%). These findings indicate that empirical whole-brain dynamics most clearly distinguished AD^+^ participants from both HC^−^ and MCI^+^ participants. The inclusion of information capability and susceptibility produced comparatively small and metric-dependent changes in classification performance. For HC^−^ vs. AD^+^, the combined specification achieved an AUC of 83.98 ± 7.25% and a balanced accuracy of 75.02 ± 8.46%, corresponding to a slight reduction in AUC but a modest increase in balanced accuracy relative to the empirical-feature model. A similar pattern was observed for MCI^+^ vs. AD^+^: AUC changed from 85.18 ± 10.73% to 84.58 ± 10.57%, whereas balanced accuracy increased from 66.37 ± 12.59% to 68.22 ± 11.15%. For HC^−^ vs. MCI^+^, adding the model-based measures produced a small increase in AUC, from 61.86 ± 9.59% to 63.18 ± 6.23%, but balanced accuracy decreased from 58.41 ± 12.86% to 55.26 ± 7.46%. Performance for HC^−^ vs. HC^+^ remained essentially unchanged (AUC: 63.07 ± 12.25% vs. 64.34 ± 11.05%; balanced accuracy: 59.45 ± 10.15% vs. 59.58 ± 12.74%).

To assess whether classification performance depended on classifier choice, we repeated the analyses using linear and polynomial-kernel support vector machines (SVMs). No alternative classifier consistently out-performed logistic regression across feature specifications, diagnostic contrasts, and performance metrics. Although linear SVM improved discrimination between HC and MCI+ and polynomial SVM improved performance for the combined HC versus AD+ classification, SVM performance was lower in several other contrasts (Table S3). These results support the use of logistic regression as a parsimonious primary model while indicating that the principal stage-related classification pattern was broadly preserved across classifiers.

SHAP analyses were then used to examine which dynamical features contributed most consistently to classification across the 40 resampling iterations (Fig. 5). In the turbulent dynamics features only models information flow at the largest spatial scales emerged as the leading feature for both contrasts involving AD^+^: information flow at *λ* = 0.03 had the highest mean absolute SHAP value for HC^−^ vs. AD^+^ and was selected in 67.5% of iterations, whereas information flow at *λ* = 0.01 was the leading feature for MCI^+^ vs. AD^+^ and was selected in 92.5% of iterations. Information flow at *λ* = 0.01 remained the highest-ranked feature for MCI^+^ vs. AD^+^ in the combined turbulent dynamics and perturbation features model, with a non-zero SHAP contribution in 87.5% of iterations (Fig. S3).

### Turbulent dynamics is spatially aligned with cortical gradients of AD-risk gene expression

Next, to explore the spatial alignment between turbulent dynamics and genetic risk factors for AD, we compared spatial patterns of cortical gene expression with the cortical distribution of group differences in turbulent dynamics. We selected 15 a priori genes from the loci compiled by the AD Sequencing Project Gene Verification Committee and previous studies (Sepulcre et al. 2018), focusing on genes implicated in the A*β* pathway (Hampel et al. 2021). Healthy reference expression profiles were obtained from microarray data in the Allen Human Brain Atlas (AHBA). Gene-expression data were processed using *abagen*, mapped to the 1,000-node Schaefer parcellation, and projected together with the turbulence-difference maps onto the fsaverage-41k surface using *neuromaps*. The selected genes were classified into three categories according to their roles in A*β* homeostasis (Fig. 6a): expression (five genes), trafficking (three genes), and degradation (seven genes).

**Figure 6.**
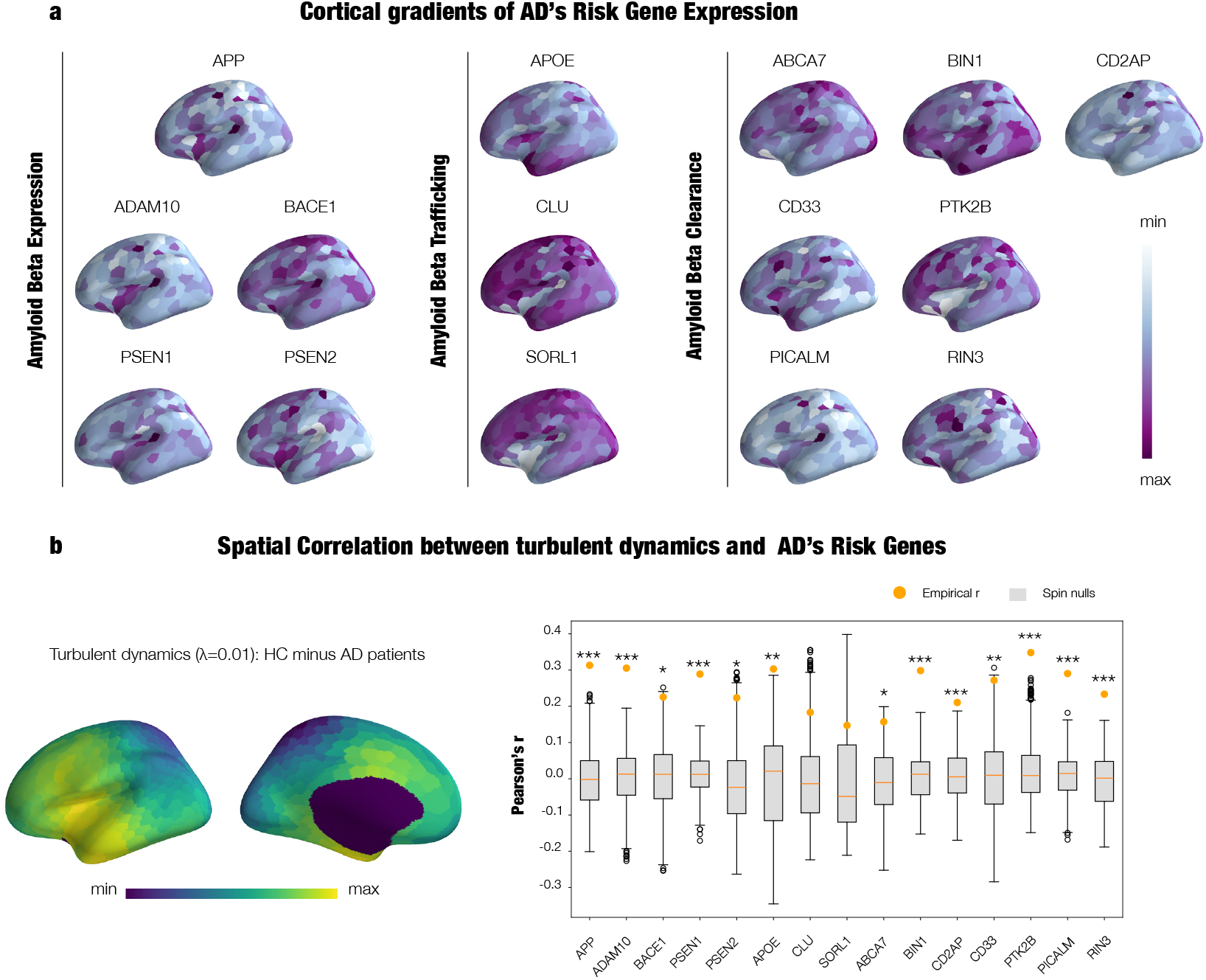
Spatial alignment between turbulent dynamics and Alzheimer’s disease-risk gene expression. (a) Cortical expression maps for 15 genes implicated in amyloid-*β* expression, trafficking, and clearance. Gene-expression values were obtained from the Allen Human Brain Atlas, mapped to the Schaefer 1,000-node parcellation, and projected onto the fsaverage-41k cortical surface. (b) Spatial correlation between the absolute HC^*−*^–AD^+^ group-mean difference in turbulent dynamics at *λ* = 0.01 and each cortical gene-expression map. Orange points show empirical Pearson correlation coefficients and grey boxplots show null distributions generated using 1,000 Alexander–Bloch spin rotations. Asterisks indicate nominal spin-test significance (^∗^*p*spin *<* 0.05, ^*∗∗*^*p*spin *<* 0.01, ^*∗∗∗*^*p*spin *<* 0.001).

Motivated by the whole-brain turbulence results, we restricted the gene-expression analysis to the HC^−^ vs. AD^+^ and MCI^+^ vs. AD^+^ contrasts, as these were the two stage comparisons that showed significant differences in turbulent dynamics at *λ* = 0.01 after FDR correction. We examined the spatial correlations between cortical gene expression and the absolute group-mean turbulence-difference maps for these contrasts (Fig. 6b, Fig. S4, and Table S4).

For the HC^−^ vs. AD^+^ contrast, the strongest spatial correspondence was observed for PTK2B (*r* = 0.348, *p*_spin_ *<* 0.001), followed by APP (*r* = 0.313, *p*_spin_ *<* 0.001), ADAM10 (*r* = 0.305, *p*_spin_ *<* 0.001), and APOE (*r* = 0.303, *p*_spin_ = 0.010). Thirteen of the 15 genes showed significant spatial associations, whereas CLU and SORL1 did not. The MCI^+^ vs. AD^+^ contrast showed a partly different gene-expression profile. The strongest correspondence was observed for CLU (*r* = 0.500, *p*_spin_ *<* 0.001), followed by CD33 (*r* = 0.418, *p*_spin_ *<* 0.001), PSEN2 (*r* = 0.413, *p*_spin_ *<* 0.001), and APOE (*r* = 0.395, *p*_spin_ = 0.012). Significant spatial associations were also observed for BACE1, RIN3, APP, and BIN1 (all *p*_spin_ *<* 0.01). Notably, APOE was the only gene ranked among the four strongest spatial correlations in both comparisons.

### Progressive decoupling between the E:I balance and turbulent dynamics maps across the AD continuum

Finally, to examine whether the topology of large-scale turbulent dynamics was related to the regional distribution of excitation and inhibition balance, we generated a normative transcriptomic E:I map from Allen Human Brain Atlas data following previous work (Deco et al. 2021) (see Methods and Fig.S5).

All four groups showed positive spatial alignment between turbulence and the E:I balance map, but the magnitude of this alignment progressively decreased across disease stages (Fig.7). Mean empirical correlations were *r* = 0.756 ± 0.079 for HC −, *r* = 0.744 ± 0.111 for HC+, *r* = 0.673 ± 0.163 for MCI+, and *r* = 0.650 ± 0.165 for AD+. In the covariate-adjusted ordered-stage model, Fisher-transformed coupling decreased significantly from HC through HC+, MCI+, and AD+ (*β*_stage_ = −0.056, HC3 95% CI [−0.091, − 0.021], *p*_perm_ = −0.0022). Relative to HC−, coupling was significantly reduced in MCI+ (*β* = −0.142, *p*_FDR_ = 0.0159) and AD+ (*β* = −0.144, *p*_FDR_ = 0.0159), but not in HC+ (*β* = −0.019, *p*_FDR_ = 0.6965).

The spatially normalised analysis produced the same stage-related pattern. Mean spin-normalised coupling was *z*_spin_ = 7.58 ± 3.46 for HC−, 6.88 ± 2.94 for HC+, 5.93 ± 2.91 for MCI+, and 5.02 ± 2.89 for AD+.

The ordered-stage effect remained significant after spatial normalisation (*β*_stage_ = −0.712, HC3 95% CI [−1.222, −0.203], *p*_perm_ = 0.0047). Compared with HC−, spin-normalised coupling was reduced in MCI+ (*β* = −1.558, *p*_FDR_ = 0.0426) and AD+ (*β* = −2.039, *p*_FDR_ = −0.0345), whereas HC+ did not differ significantly from HC (*β* = −0.613, *p*_FDR_ = 0.3710). These findings suggest a progressive reduction in the spatial coupling between large-scale turbulent dynamics and the E:I balance maps.

## DISCUSSION

In the present study, we combined multiscale turbulent dynamics measures with in silico responses to external perturbations using whole-brain Hopf modelling to investigate disruptions across the AD continuum. We used whole-brain models with generative effective connectivity at both group- and subject-level, with the latter enabling the investigation of how perturbational responsiveness could be related to AT(N) biomarkers and cognitive impairment. In an exploratory analysis, we also aimed to examine the spatial relationship between turbulent dynamics and cortical patterns of AD-risk gene expression as well as the E:I balance.

Our first main finding is that AD is characterised by a scale-dependent reorganisation of turbulent dynamics. We observed reduced turbulence at large spatial scales together with increased dynamics at intermediate–small scales, accompanied by decreases in information flow and transfer. Crucially, all three large-scale turbulence and information measures were significantly associated with MoCA scores after adjustment for age, sex, and education with information flow accounting for the largest descriptive proportion of the dynamics-related explained variance. This pattern suggests impaired long-range integration coupled with dysregulated local interactions.

At the network level, the DMN contained the largest absolute number of nodes among those showing the strongest statistical effects, consistent with the prominent involvement of this network in AD. Notably, early amyloid deposition is thought to preferentially target medial parietal and frontal association cortices —core hubs of the DMN— placing this network at the centre of the initial pathological cascade (Palmqvist et al. 2017).

**Figure 7.**
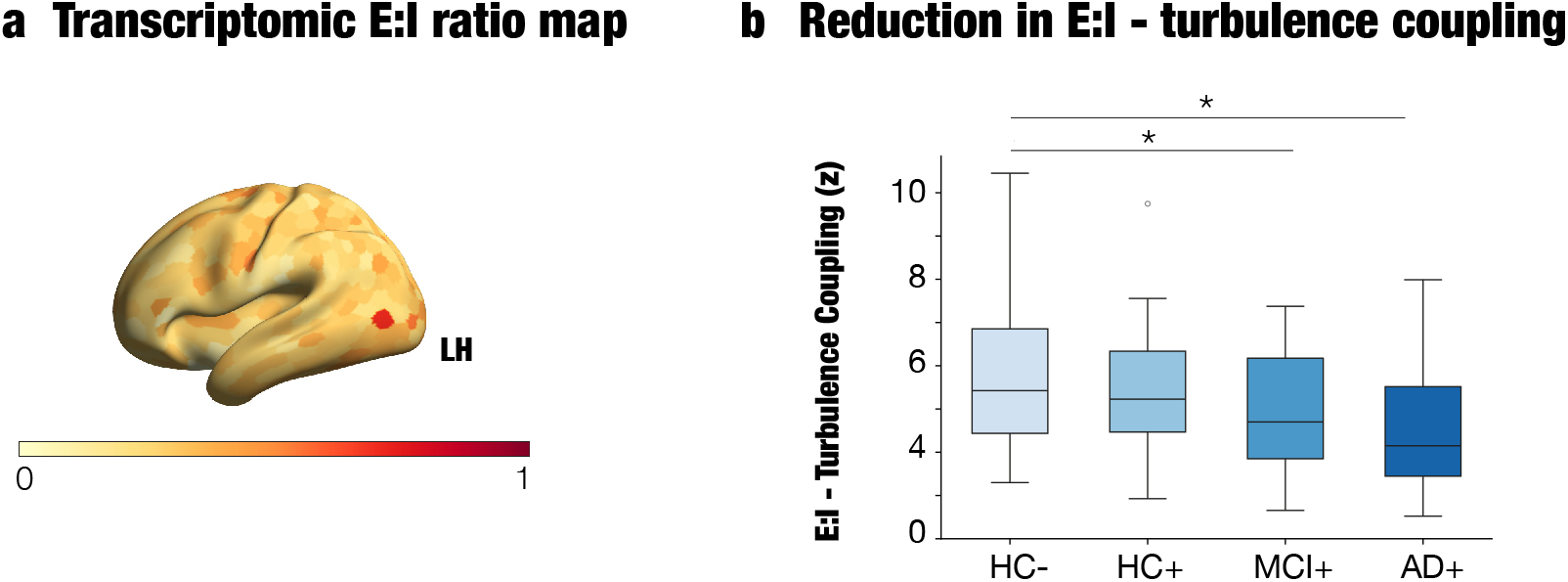
Progressive reduction in the spatial alignment between the excitation–inhibition balance and large-scale turbulence maps. (a) Cortical excitation-to-inhibition (E:I) ratio derived from Allen Human Brain Atlas gene-expression data. The map represents the ratio between excitatory glutamatergic receptor genes and inhibitory GABAergic receptor genes (see Methods). (b) Participant-level spatial correspondence between the E:I ratio map and turbulence at *λ* = 0.01. Coupling decreased significantly across the ordered HC^*−*^, HC^+^, MCI^+^, and AD^+^ stages (*β*_stage_ = −0.712, permutation *p* = 0.0047), after adjustment for age, sex, and education using 10,000 Freedman–Lane permutations. Asterisks indicate planned comparisons with HC^*−*^ that remained significant after FDR correction (^∗^*p*FDR *<* 0.05).

The current findings align with our recent results showing a disruption of the hierarchical brain organization in the AD continuum with changes in trophic levels in the DMN significantly associated with cognitive impairment (Montaña-Valverde et al. 2026). The breakdown of this architecture might help to explain the alterations in turbulent dynamics, information flow and information transfer measures observed in the current study.

Prior work has also shown reduced DMN connectivity in rsfMRI (Agosta et al. 2012, Koch et al. 2010, Qi et al. 2010, Sorg et al. 2007), impaired task-related deactivation (Celone et al. 2006, Petrella et al. 2007, Rombouts et al. 2009), and spatial overlap with amyloid accumulation in preclinical stages (Hedden et al. 2009). DMN connectivity is also altered in cognitively normal individuals with elevated amyloid burden (Hedden et al. 2009, Oh et al. 2011, Sheline et al. 2010b), in relation to tau-related CSF biomarkers (Li et al. 2013, Molinuevo et al. 2013, Wang et al. 2013), and even in amyloid-free APOE4 carriers (Sheline et al. 2010a), suggesting that DMN dysfunction may precede overt pathology. Our results extend this literature by showing that DMN vulnerability is expressed not only in static connectivity changes, but also in the breakdown of turbulent dynamics at different spatial scales.

At the biomarker level, large-scale turbulent dynamics were significantly associated with all three components of the AT(N) framework: lower turbulence was associated with greater amyloid burden and tau deposition, whereas higher turbulence was associated with larger bilateral hippocampal volume.

Our second finding was that responses to in silico perturbations of whole-brain models —information capability and susceptibility— exhibit a progressive reduction across all AD stages, capturing both early transitions from A*β*-to A*β*+ healthy controls and later stages of clinical impairment. Beyond their sensitivity to disease staging, both measures were significantly associated with proteinopathy, including both neocortical A*β* PET burden and regional tau PET in areas across different Braak stages. These results highlight the importance of perturbation-based measures as complementary neuroimaging markers for AD staging incorporating A*β* status.

Our third main finding was that model-free measures discriminated comparisons involving AD^+^ with relatively high ROC–AUC, whereas classification was more modest for preclinical and earlier-stage comparisons. Adding information capability and susceptibility produced comparatively small and metric-dependent changes rather than a consistent improvement across classifiers. Thus, the current findings do not establish a general performance advantage for the combined specification, although the model-based measures may provide complementary information in particular stage comparisons. Feature-importance analyses further indicated that large-scale information flow was a prominent classifier feature in both comparisons involving AD^+^, consistent with its association with cognitive performance.

The present study builds on prior whole-brain modelling work demonstrating that AD is characterised by progressive reductions in large-scale coherence and metastability, reflecting a displacement of brain dynamics from an optimal operating regime (Córdova-Palomera et al. 2017, Demirtaş et al. 2017). In that framework, our group previously reported that CSF A*β*_1–42_ was primarily associated with the distinction between healthy and clinical groups, whereas CSF t-tau and p-tau showed more robust associations with metastability across the disease spectrum, suggesting a tighter coupling between tau pathology and progressive disruptions in large-scale functional integration (Demirtaş et al. 2017).

The present results extend the abovementioned findings in several key respects. First, by using PET-based biomarkers, we situate whole-brain dynamical measures within the full AT(N) framework using anatomically resolved indices of amyloid burden, tau deposition, and neurodegeneration. Second, the use of tau-PET aggregate values corresponding to successive Braak stages reveals that reduced responsiveness to perturbation is associated not only overall tau burden but with its hierarchical propagation from mesial temporal to temporoparietal regions, an effect that cannot be resolved using CSF measures alone. Furthermore, we showed that information flow, a measure of brain dynamics that can only be calculated when multiple spatial scales are considered, explained most of the variance in cognitive impairment.

Our fourth main finding is that the spatial distribution of large-scale dynamical alterations showed correspondence with cortical gradients of AD-risk gene expression. For the HC^−^ vs. AD^+^ contrast, the strongest spatial correlations at *λ* = 0.01 were observed for *PTK2B, APP, ADAM10*, and *APOE* (*r* = 0.303, *p*_spin_ = 0.010). A partly different profile was observed for MCI^+^ vs. AD^+^, for which the strongest correlations involved *CLU, CD33, PSEN2*, and *APOE*. Notably, *APOE* was the only gene ranked among the four strongest spatial correlations in both contrasts, suggesting a spatial relationship that may extend across the transition from healthy ageing to AD as well as between symptomatic disease stages. The recurring correspondence with *APOE*, a major genetic determinant of AD susceptibility, is also consistent with evidence linking the *ε*4 allele to increased A*β* deposition, earlier disease onset, tau-related processes, and network-level propagation (Dincer et al. 2022, Montal et al. 2022, Yamazaki et al. 2019).

Our fifth main finding was a progressive reduction in the spatial correspondence between large-scale turbulent dynamics and the normative transcriptomic map of excitation and inhibition. Although subject-level turbulence maps remained positively coupled to the E:I balance in every group, this correspondence decreased across the ordered HC−, HC+, MCI+, and AD+ stages. Importantly, the ordered-stage effect remained significant after controlling for age, sex, and education and after normalising each participant’s empirical correlation against spatially constrained spin nulls. The result therefore cannot be explained solely by the broad spatial smoothness or autocorrelation shared by cortical maps. Instead, it suggests that AD progression is accompanied by an increasing reorganisation of large-scale dynamics away from the cortical pattern normally associated with regional variation in excitatory and inhibitory receptor expression.

This finding converges with evidence that AD is associated with disturbances in E:I regulation, including neuronal hyperactivity near amyloid plaques, inhibitory dysfunction, aberrant synchronisation, and shifts towards more supercritical dynamical regimes (Busche et al., 2008; Busche and Konnerth, 2015; Palop and Mucke, 2016; Maestú et al., 2021; Javed et al., 2025). It is also consistent with our previous whole-brain modelling work showing that increasing amyloid-related scaling can alter the simulated E:I ratio (Patow et al., 2023). The present analysis extends these observations by demonstrating that the relationship can be detected at the level of individual spatial maps: participants at more advanced disease stages showed progressively weaker alignment between empirical turbulent dynamics and the E:I balance map.

Several limitations of the present study should be acknowledged. First, although we leveraged a multimodal dataset combining rsfMRI, structural MRI, PET imaging, and computational modelling in a moderately sized cohort from ADNI3, the cross-sectional nature of our analyses precludes strong inferences about the temporal dynamics of turbulent alterations across the AD continuum. Longitudinal data will be essential for validation. Second, our analysis relied on a single parcellation scheme (Schaefer 1000-node atlas) to compute turbulent dynamics. While this fine-grained atlas ensured comparability with prior work with turbulent dynamics, replication across multiple parcellations would strengthen generalisability. Third, the canonical structural connectivity was based on an independent HCP sample as it has produced robust and reproducible results with whole-brain Hopf models in different neuropsychiatric (Escrichs et al. 2024) and neurological (Martínez-Molina et al. 2024) populations. Nonetheless, it is important to note that we used GEC for whole-brain modelling, which is subject-specific as it’s fitted to each individual’s FC. It also is important to acknowledge that while our model-based measures are highly sensitive to disease stage at the group level, they showed reduced sensitivity when examined at the subject-level. We believe this might be a reflection of the challenges in mapping complex dynamics to individual diagnostic status using current generative models. Fourth, because the stage-specific samples were modest, particularly for AD^+^, regressions relating dynamical measures to AT(N) biomarkers and cognition were performed in the pooled cohort to preserve statistical power. Consequently, the observed associations may partly reflect between-stage differences and should not be interpreted as evidence of participant-level relationships within individual disease stages. In addition, transcriptomic associations were based on microarray-derived gene expression from the AHBA, which is limited to a small number of healthy donors. Finally, our results for the altered coupling with the E:I balance across the AD continuum are not a direct measurement of participant-specific E:I balance. Future studies combining longitudinal functional imaging with participant-level measures sensitive to E:I physiology will be needed to clarify the molecular and physiological mechanisms underlying this progressive decoupling.

In sum, we demonstrate that AD is characterised by convergent alterations in model-free and model-based measures, with the latter showing robust and consistent differences across all disease stages. These findings position whole-brain model-based measures as promising neuroimaging biomarkers that capture stage-dependent and continuous aspects of AD progression. In parallel, model-free turbulent dynamics reveal a scale-dependent reorganisation, linking impaired large-scale coordination with dysregulated local activity. Together, these approaches provide a unified systems-level account in which proteinopathy, neurodegeneration, and largescale brain dynamics are tightly coupled. The strong associations with AT(N) biomarkers, particularly neurodegeneration, and the convergence with transcriptomic gradients of AD-risk genes further support the biological relevance of brain dynamics measures. Importantly, we also found an association between turbulent dynamics and cognitive impairment. Moreover, the decoupling between E:I balance and turbulent dynamics points to a potential mechanism bridging microscopic and macroscale alterations. Thus present results highlight the importance of model-based and model-free measures of brain dynamics as valuable complementary neuroimaging markers in AD progression.

## MATERIALS AND METHODS

### ADNI Participants

Data for this study were obtained from the Alzheimer’s Disease Neuroimaging Initiative (ADNI) database (https://adni.loni.usc.edu/). ADNI was established in 2003 as a public–private partnership under the leadership of Michael W. Weiner, MD. Initially, the initiative aimed to investigate how longitudinal MRI, PET, biological biomarkers, and clinical and neuropsychological assessments could be integrated to characterise the progression of mild cognitive impairment and early Alzheimer’s disease. Its current objectives include biomarker validation for clinical trials, improving cohort diversity and generalisability, and supporting research into Alzheimer’s disease diagnosis and progression through data sharing.

151 participants were included from the ADNI3 database on the basis of the availability of clinical data, baseline [18F] florbetapir/florbetaben amyloid-PET, 3-T resting-state fMRI, and T1-weighted MRI. All clinical data had to be obtained within a time frame of one month with respect to baseline neuroimaging. A participant’s cognitive status was determined by ADNI as Healthy Control (HC) [Mini-Mental State Examination (MMSE) ≥ 24, Clinical Dementia Rating (CDR) = 0, nondepressed], Mild Cognitive Impairment (MCI) [MMSE ≥ 24, CDR = 0.5, objective memory impairment on education-adjusted Wechsler Memory Scale II, preserved activities of daily living], and Alzheimer’s Disease (AD) [MMSE 20 ≤ MMSE ≤ 26, CDR ≥ 0.5, National Institute of Neurological and Communicative Disorders and Stroke/Alzheimer’s Disease and Related Disorders Association criteria for probable AD].Amyloid status was determined using a global cut-off (see PET processing). The combined staging included healthy controls A*β*^−^ (HC^−^) and A*β*^−^ (HC^+^), MCI A*β*^+^ (MCI^+^), and AD A*β*^+^ (AD^+^). All study procedures were conducted in accordance with the Declaration of Helsinki, and ethical approval was obtained by ADNI investigators. All study participants provided written informed consent. For more details,please see https://adni.loni.usc.edu/wp-content/themes/freshnews-dev-v2/documents/clinical/ADNI3_Protocol.pdf.

### Neuropsychological testing

We selected cognitive assessments based on their clinical relevance and widespread use in dementia research, prioritising those available for the largest number of participants in our dataset. These included the previously mentioned MMSE (Folstein et al. 1975) as well as the Alzheimer’s Disease Assessment Scale–Cognitive (ADAS-Cog-13) 13-item version (Rosen et al. 1984), and the Montreal Cognitive Assessment (MoCA) (Nasreddine et al. 2005). This set of measures was chosen to capture a broad range of cognitive domains, including global cognition, memory, language, executive functioning, and functional abilities relevant to daily living. For each participant, we included the cognitive assessments corresponding to the same study phase as the MRI acquisition (ADNI3) and available for the same individuals in the demographics dataset. Only baseline or screening visits (e.g., bl, sc) were retained to ensure that the cognitive assessments were conducted in close temporal proximity to the corresponding imaging session. In cases where multiple assessments were available for a given participant, the earliest visit was selected. Additional details on the cognitive measures are provided in the ADNI protocols documentation https://adni.loni.usc.edu/help-faqs/adni-documentation/.

### MRI acquisition

Structural MRI and fMRI were acquired using 3T Siemens and GE scanners. T1-weighted structural scans were collected using either an MPRAGE (TR = 2300 ms; voxel size = 1 × 1×1 mm) or IRFSPGR sequence (TR = 7.3 ms; voxel size = 1 × 1×1 mm). For parameter details, see https://adni.loni.usc.edu/wp-content/uploads/2017/07/ADNI3-MRI-protocols.pdf. Resting-state fMRI (eyes open) was performed using a 3D echo-planar imaging (EPI) sequence with 197 fMRI volumes per patient (TR/TE =3000/30 ms; flip angle =90°; voxel size =3.4 × 3.4 × 3.4 mm). PET data were assessed after intravenous injection of 18F-labeled tracers (flortaucipir: 6× 5-min time frames, 75 to 105 min after injection; florbetapir: 4× 5-min time frames, 50 to 70 min after injection; florbetaben: 4× 5-min time frames, 90 to 110 min after injection). For more information, see https://adni.loni.usc.edu/datasamples/adni-data/neuroimaging/pet/.

### Structural MRI preprocessing

Structural MRI data were processed using the voxel-based morphometry (VBM) pipeline implemented in SPM12 (https://www.fil.ion.ucl.ac.uk/spm/). Prior to preprocessing, all images were manually reoriented to the anterior commissure to ensure consistent alignment. Segmentation of T1-weighted scans into gray matter (GM), white matter (WM), and cerebrospinal fluid (CSF) compartments was performed using the unified segmentation approach (Ashburner and Friston 2005). The baseline GM, WM and CSF segmentations were then used to generate a group average template using the diffeomorphic warping “Shoot” algorithm in SPM12 (Ashburner and Friston, 2011), which provides high-dimensional alignment of individual gray matter probability maps to a population-specific template. The segmentations were then warped using the resulting deformation fields, modulated with the Jacobian determinant data and smoothed with a 6mm full width half maximum (FWHM) Gaussian kernel.Total Intracranial Volume (TIV) was computed in SPM12 using the Utilities → Tissue Volumes module applied to each participant’s native-space segmentation. Following standard practice, we summed three tissue classes (GM, WM, CSF) within the SPM intracranial mask to obtain whole-brain intracranial volume in millilitres. ROI-based gray matter volumes (GMV) were extracted from the hippocampus, as defined by the Brainnetome atlas (Fan et al. 2016).

### rsfMRI preprocessing

We followed the preprocessing pipeline described in our previous work (Martínez-Molina et al. 2024), which is based on the standard volume-based pipeline implemented within the Matlab-based CONN toolbox (http://www.nitrc.org/projects/conn), version 22v2407. The following steps are included in the standard CONN pipeline: functional realignment and motion correction; slice-timing correction to account for differences in time of acquisition between slices; identification of outlier scans for subsequent regression by means of the quality assurance/artifact rejection software Artifact Detection Toolbox (art;http://www.nitrc.org/projects/artifact_detect); coregistration with each participant’s high-resolution T1-weighted image; spatial normalisation to MNI-152 standard space with 2mm isotropic resampling resolution and smoothing with a Gaussian kernel of 8mm FWHM.

Denoising also followed the same procedure as in our previous work: to reduce noise due to cardiac and motion artifacts we applied the anatomical CompCor method (Behzadi et al. 2007)(also implemented within the CONN toolbox), by regressing out of the functional data the first five principal components attributable to each individual’s white matter signal; the first five components attributable to individual cerebrospinal fluid (CSF) signal; six subject-specific realignment parameters (three translations and three rotations) as well as their first-order temporal derivatives; the artifacts identified by art; and main effect of scanning condition. Finally, after linear detrending, each individual’s denoised BOLD signal timeseries were band-pass filtered in the 0.008-0.08 Hz range, to eliminate both low-frequency drift effects and high-frequency noise.

### PET preprocessing

Downloaded PET data had previously been co-registered to the first scan and averaged (stage 2), by the PET Core Group and scans labeled with bad quality were discarded. Preprocessing was performed in the Statistical Parametric Mapping (SPM12,https://www.fil.ion.ucl.ac.uk/spm/software/spm12/) including co-registration of each PET image to the native T1-weighted MRI, which was then segmented to obtain the deformation fields used for the normalization of the PET scans to MNI-152 standard space with 2mm isotropic resampling resolution. PET scans were smoothed with a Gaussian kernel of 8mm FWHM based on each scan’s metadata and resolution table. Partial volume correction was applied after the co-registration step using the Müller-Gärtner algorithm implemented in the PETPVC toolbox (Thomas et al. 2016). All PET scans for amyloid tracers, Florbetapir (FBP) and Florbetaben (FBB), were converted to Standardized Uptake Value Ratios (SUVRs) using the whole cerebellum as reference (Bullich et al. 2017, Cho et al. 2020). Both FBP and FBB SUVRs were converted to Centiloid (CL) units using specific calibration equations (FBB: CL = 94.082 × SUVR – 94.772; FBP: CL = 110.982 × SUVR – 116.172), following the GAAIN Centiloid Project Level-1 and Level-2 procedures (Klunk et al., 2015). Amyloid status was determined with a global cutoff of 24 centiloids (equivalent to SUVR 1.11 FBP / 1.08 FBB) (Navitsky et al. 2018).

### Turbulence

We investigated the turbulent brain dynamics by measuring the information processing across spacetime scales with the local Kuramoto order parameter. The degree of synchronisation of a particular brain region (*n*) is characterized as a function of space 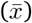 and time (*t*) at a given scale (*λ*) (Fig. 1a–b). The parameter *λ* defines the extent of the spatial distances (i.e., *λ* scales) where the local synchronisation is assessed. High *λ* values represent short distances in the brain, while small *λ* values represent long distances in the brain (e.g. *λ*=0.01 (approximately 100 mm) and *λ*=0.3 (approximately 3 mm).

#### Amplitude turbulence

We measure amplitude turbulence by first defining the local Kuramoto order parameter and then taking the standard deviation of the modulus across time and space. First, we define the amplitude turbulence, *R*_*λ*_ (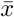, *t*), as the modulus of the local Kuramoto order parameter for a given brain region as a function of the time at each spatial scale *λ*:

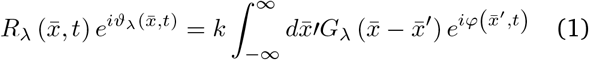

where 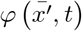 denotes the phases of the BOLD signal data, G_*λ*_ refers to the local weighting kernel 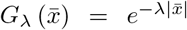 denotes the normalisation factor 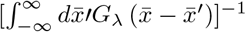, and λ denotes the spatial scaling. Therefore, local levels of synchronisation at a certain λ, as a function of space 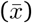 and time (t) are defined by R_λ_.

We then measure the amplitude turbulence, D, as the standard deviation across nodes and time of the modulus of the local Kuramoto order parameter (R_λ_):

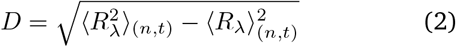

where the brackets <>_(*n,t*)_ denotes averages across nodes and time.

#### Information cascade flow

The information cascade flow indicates how the information travels from one scale (*λ*) to a lower scale (*λ* − Δ*λ*, where Δ*λ* corresponds to a scale step) in successive time steps (*t* and *t* + Δ*t*). In this regard, the information cascade flow captures the information transfer across scales, and it is calculated as the temporal correlation between the local Kuramoto order parameter in two successive scales and times as:

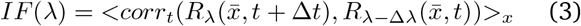

where the brackets <>_*x,t*_ indicate averages across time and nodes.

#### Information transfer

The information transfer captures how information travels across space at each λ scale. It is calculated as the slope of a linear fitting, in the log-log scale, of the temporal correlation between the local Kuramoto order parameter of two nodes at each scale as a function of its Euclidean distance (*r*) in the inertial subrange

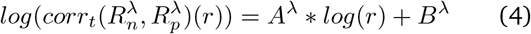

where *A* and *B* are the fitting parameters for the *λ* scale, and *r* denotes the distance in the brain. The negative slope (*A*^*λ*^) represents the transfer in the spatial direction *r* of the information in terms of temporal correlation of the local synchronisation level. Thus, when the slope is more pronounced, the information travels short distances, while a flatter slope indicates that information is transferred over long distances.

#### Local node-level turbulence

The node-level turbulence, NLT, defines the variability of the local synchronisation of the nodes and is computed as the standard deviation across time of the local Kuramoto order parameter as follows:

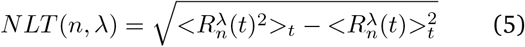

where brackets <>_*t*_ denote average across time. Here, we estimated the discrete version of the node-level Kuramoto order parameter, with modulus R and phase *v*, which represents a spatial average of the complex phase factor of the local oscillators weighted by the coupling as:

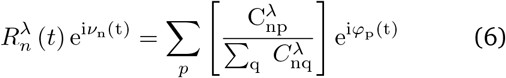

where φ_p_(t) represents the phases of the resting-state BOLD signal data, and 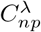 refers to the local geometric weighting kernel between nodes *n* and *p*:

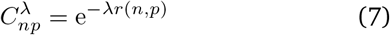

where *r*(*n, p*) is the Euclidean distance between nodes *n* and *p* in MNI space, and *λ* controls the spatial extent of the distance-decay kernel. This geometric kernel was used to calculate turbulence observables and is distinct from the fitted GEC matrix used to couple the Hopf oscillators.

### Data harmonisation

We used ComBat, a widely adopted and reliable harmonisation tool that has demonstrated high accuracy in eliminating site effects (Bell et al. 2022, Yu et al. 2018), outperforming alternative methods such as the traveling-subject approach (Roffet et al. 2022). ComBat was originally developed by Johnson et al. (2007) as a batch-effect correction tool for genomics data, and was later adapted for neuroimaging applications by Fortin et al. (2018) to regress out site-induced variance in MRI datasets. ComBat operates within an empirical Bayes framework, fitting a set of biological covariates to the data and then applying a linear shift-and-scale transformation to the model residuals to align them with a reference distribution. The covariate effects are subsequently reintroduced, yielding an adjusted dataset for each site while preserving the influence of biological variables. ComBat uses a multivariate linear mixed-effects regression with terms for biological variables and scanner to model imaging measurements as follows:

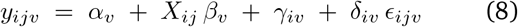

where *Y*_*ijv*_ represents the brain measure (e.g., turbulent dynamics measures) at site *i* and participant *j* for feature *v*. The term *α*_*v*_ denotes the average of the measure across participants, *X*_*j*_ is the design matrix for covariates of interest, and *β*_*v*_ is the vector of regression coefficients corresponding to *X*_*j*_. The parameters *γ*_*iv*_ and *δ*_*iv*_ represent the additive and multiplicative site effects for site *i* and feature *v*, respectively, and *ϵ*_*ijv*_ denotes an error term assumed to follow a normal distribution with zero mean and variance 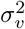. The ComBat-harmonised brain measures are then defined as:

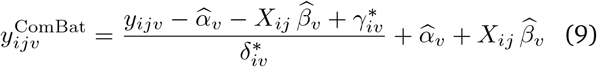

where 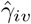 and 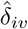 are the empirical Bayes estimates of γ_*iv*_ and δ_*iv*_, respectively.

Data harmonisation was implemented in Python using the `harmonizationLearn` and `harmonizationApply` functions from the `neuroHarmonize` package (Pomponio et al. 2020), with covariates age, gender, education, and diagnosis. Since harmonisation methods rely on data from multiple sites for effective calibration, 6 subjects with unique sites were excluded from the original sample resulting in a total sample size N=145.

### Machine learning analysis

We implemented binary logistic-regression classifiers for four prespecified comparisons: HC^*−*^ vs. HC^+^, HC^*−*^ vs. MCI^+^, HC^*−*^ vs. AD^+^, and MCI^+^ vs. AD^+^. Each comparison was evaluated using two feature specifications. The empirical-dynamics specification included amplitude turbulence across 10 spatial scales, information flow across nine adjacent scale pairs, information transfer across 10 spatial scales, information cascade, metastability, and the global Kuramoto order parameter. The combined specification incorporated the participant-level information capability and susceptibility estimates in addition to the eligible empirical-dynamics features. Information capability and susceptibility were always retained in the combined models.

For each comparison and feature specification, we performed 40 repeated stratified random partitions of the data into 75% training and 25% held-out test sets. All preprocessing, feature selection, hyperparameter optimisation, and model fitting were restricted to the training data. Features were standardised using parameters estimated from the corresponding training data and the same transformation was subsequently applied to the held-out test set.

Within each outer training set, minimum-redundancy maximum-relevance (mRMR) feature selection was used to rank the empirical dynamical features. The mRMR ranking was estimated from the complete outer training set and was not exposed to the held-out test data. Inner leave-one-out cross-validation (LOOCV) was then used to jointly optimise the number of retained features, the inverse regularisation strength, and the regularisation penalty. The number of retained features was evaluated from 5 to 15, the inverse regularisation parameter was evaluated at *C* = {0.01, 0.1, 1, 5, 10, 50, 100, 500}, and both L1 and L2 penalties were considered. Hyperparameters were selected by maximising the inner-LOOCV ROC–AUC. Logistic-regression models used class-balanced weights, the liblinear solver, and a maximum of 1,000 optimisation iterations.

After hyperparameter selection, mRMR was reapplied to the complete outer training set using the selected number of features, and the final logistic-regression model was fitted to the complete training data. A decision threshold was selected from the training data by maximising Youden’s *J* statistic and was subsequently applied without modification to the held-out test set. Performance was quantified using ROC–AUC, balanced accuracy, sensitivity, and specificity. Metrics were aggregated across the 40 outer repetitions and are reported as mean ± SD.

Feature importance was examined using SHapley Additive exPlanations (SHAP). Kernel SHAP values were calculated for the selected features in the held-out test sample from each repetition, using predicted probability for the positive class as the model output. Feature importance was summarised as the mean absolute SHAP value. To distinguish the magnitude of a feature’s contribution from the consistency with which it was retained, we additionally calculated its selection frequency across the 40 repetitions. Aggregate SHAP importance values therefore reflect both the magnitude of a selected feature’s contribution and the frequency with which it entered the fitted classifiers.

Classification was repeated using linear- and polynomial-kernel support vector machines (SVMs), while retaining the same 40 repeated 75/25 train–test partitions and nested preprocessing pipeline used for logistic regression. Feature standardisation and mRMR selection were performed using the training data only. Hyperparameters were optimised by inner leave-one-out cross-validation using ROC–AUC: both classifiers tested *C* ∈ {0.01, 0.1, 1, 10, 100} and 5–15 selected features, while the polynomial SVM additionally tested degrees 2 and 3 (*γ* = scale; coef0 = 1). Balanced class weights and probability estimation were used for both classifiers.

### Whole-brain Hopf model

We constructed whole-brain models with *N* = 1,000 cortical regions, in which the local dynamics of each region were represented by the normal form of a supercritical Hopf bifurcation (a Stuart–Landau oscillator) (Deco et al. 2017, Ponce-Alvarez and Deco 2024). For an uncoupled region *n*, the dynamics were

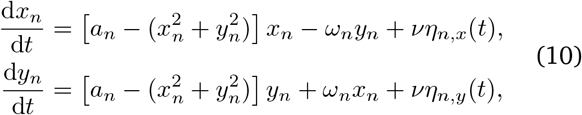

where *a*_*n*_ is the local bifurcation parameter, *ω*_*n*_ = 2*πf*_*n*_ is the intrinsic angular frequency estimated from the peak of the empirical BOLD power spectrum, and *η*_*n,x*_ and *η*_*n,y*_ are independent Gaussian white-noise processes with amplitude *v* = 0.01. For *a*_*n*_ *<* 0, the oscillator has a stable fixed point and exhibits noise-driven fluctuations; for *a*_*n*_ *>* 0, it has a stable limit cycle; and *a*_*n*_ = 0 is the bifurcation point. The unperturbed simulations used *a*_*n*_ = *−*0.02 for every region (Fig. 1e).

#### Generative effective connectivity estimation

The interregional coupling matrix used in the simulations was not the raw structural-connectivity matrix. Instead, we estimated a generative effective-connectivity (GEC) matrix that was initialized from structural connectivity and fitted to the empirical zero-lag functional connectivity and one-TR lagged normalized covariance using a pseudo-gradient descent algorith,. This procedure follows the linear approximation of the Hopf model and prior GEC implementations from the same modelling framework (Ponce-Alvarez and Deco 2024, Stikvoort et al. 2025). Let **u** = (*x*_1_, …, *x*_*N*_, *y*_1_, …, *y*_*N*_)^T^. Around the stable fixed point, the dynamics are approximated by

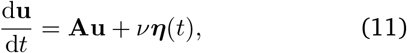

with Jacobian

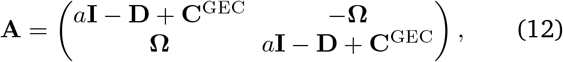

where *a* = − 0.02, **Ω** = diag(*ω*_1_, …, *ω*_*N*_), and *D*_*nn*_ =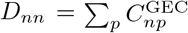. The stationary covariance **Σ** and its lagged counterpart were obtained analytically as

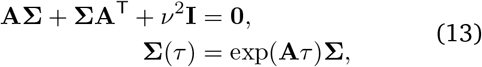

where *τ* was one repetition time (TR = 3 s). Model functional connectivity was obtained by variance-normalizing the zero-lag covariance, and the lagged statistic was obtained by variance-normalizing the upper-left *N* ×*N* block of ∑(*τ*). At fitting iteration *k*, each connection permitted by the model mask—either a non-zero structural connection or the corresponding interhemispheric parcel pair (index offset 500 in the Schaefer-1,000 ordering)—was updated according to

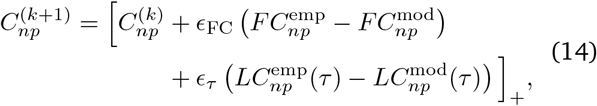

where *LC*(*τ*) denotes normalized lagged covariance and [·]_+_ truncates negative values at zero. The learning rates were *ϵ*_FC_ = 1.8 *×* 10^*−*4^ and *ϵ*_*τ*_ = 3.6 × 10^*−*4^. After each iteration, the largest GEC weight was rescaled to 0.1, and fitting continued until the combined mean-squared error for the two empirical targets converged.

#### Nonlinear whole-brain Hopf model and global coupling

Before nonlinear simulation, each fitted GEC matrix was rescaled so that its maximum weight was 0.2. The coupled whole-brain dynamics were then

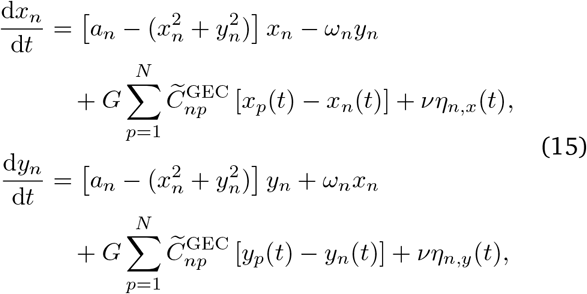

where 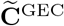 is the rescaled fitted GEC matrix and the global coupling parameter *G* scales all incoming connections equally. The complex state is *z*_*n*_(*t*) = *x*_*n*_(*t*) + i*y*_*n*_(*t*);the real component *x*_*n*_(*t*) was used as the simulated BOLD-like signal, whereas *y*_*n*_(*t*) is the latent oscillator component. The intrinsic frequencies were fixed during each simulation.

The optimal global coupling was selected by matching the distance-dependent correlation structure of the simulated and empirical BOLD signals. For a variable *u*, the second-order structure function is

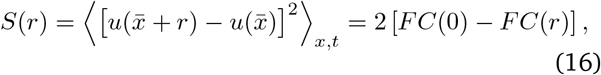

where

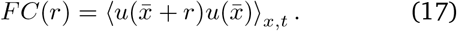

For each candidate value of G, we calculated the Euclidean discrepancy between simulated and empirical FC(r) within the prespecified inertial subrange and retained the value that minimized this error. Thus, GEC fitting estimated connection-specific weights from empirical zero- and one-TR-lagged statistics, whereas the subsequent *G* optimization set a single global scale for the nonlinear simulations.

#### Susceptibility and Information encoding capability

We used the susceptibility and information capability measures in order to study how the whole-brain model reacts to *in silico* perturbations. The susceptibility measure obtains the brain’s sensitivity to react to external stimuli. The Hopf model was perturbed for each *G* by randomly changing the local bifurcation parameter, *a*_*n*_, in the range [-0.02: 0] and was computed by measuring the modulus of the local Kuramoto order parameter as:

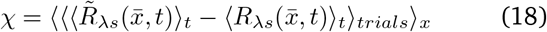

where 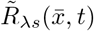 denotes the perturbed case, 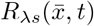 the unperturbed case, and ⟨ ⟩_*t*_ the average across time, ⟨ ⟩_*trials*_ trials, ⟨ ⟩_*x*_ and space. Similarly, the Information capability (I) captures how the external stimuli are encoded in whole-brain dynamics. This measure was defined as the standard deviation across trials of the difference between the perturbed 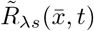 and unperturbed 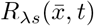 mean of the modulus of the local Kuramoto order parameter across time *t*, averaged across all brain regions *n* as follows:

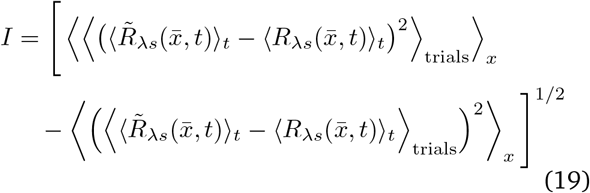

where the brackets ⟨ ⟩_*t, trials*_, and ⟨⟩_*s*_ represent the average across time, trials, and space, respectively.

### Spatial association between turbulent dynamics and gene expression

To investigate whether large-scale alterations in turbulent dynamics are associated with spatial patterns of AD-risk gene expression, we performed a surface-based spatial correlation analysis using the `neuromaps` toolbox. A voxelwise map capturing group differences in turbulent dynamics between healthy controls (HC–) and AD+ patients at the largest spatial scale (*λ* = 0.01) was used as the reference map. This map was defined in MNI152 2 mm volumetric space and subsequently projected onto the fsaverage cortical surface (41k vertex density) using the `mni152_to_fsaverage transformation`.

To account for spatial autocorrelation, we generated a null distribution of surrogate maps using the Alexander–Bloch spin permutation method as implemented in `neuromaps`. Specifically, 1000 spatially constrained rotations of the reference turbulence map were computed on the fsaverage surface, preserving its spatial covariance structure while disrupting its anatomical alignment.

We examined the spatial correspondence between the turbulence map and cortical expression patterns of a set of AD-risk genes selected based on prior genetic and transcriptomic evidence (APP, ADAM10, BACE1, PSEN1, PSEN2, APOE, CLU, SORL1, ABCA7, BIN1, CD2AP, CD33, PTK2B, PICALM, RIN3) (Sepulcre et al. 2018). Gene expression maps were obtained in MNI152 2mm space and projected to fsaverage 41k using the same transformation pipeline applied to the turbulence map.

For each gene, we computed the Pearson correlation coefficient between the turbulence map and the corresponding gene expression map using `compare_images`. Statistical significance was assessed using the precomputed spin-based null distributions, yielding a nonparametric *p*_spin_ value that reflects the proportion of surrogate correlations exceeding the empirical association.

### Construction of the transcriptomic excitation–inhibition map

A normative cortical excitation–inhibition (E:I) map was generated from post-mortem gene-expression data from the Allen Human Brain Atlas using abagen (version 0.1.3). Microarray samples from all available donors were mapped to the Schaefer 1,000-parcel, seven-network atlas in MNI152 2-mm space. Probe annotations and MNI coordinates were updated using the reannotated probes and corrected sample coordinates provided by abagen. Probes that did not exceed background intensity in at least 50% of samples were excluded. When multiple probes represented the same gene, a representative probe was selected using RNA-sequencing data, with probe selection aggregated across donors. Tissue samples were assigned to parcels using a 2-mm tolerance. For parcels without a directly assigned sample, expression was estimated from the tissue sample nearest to the parcel centroid.

No sample-wise normalization was applied. Gene-expression values were normalized across tissue samples using scaled robust sigmoid normalization and subsequently averaged across donors. Left-to-right mirroring was not performed; expression was estimated independently for all 1,000 cortical parcels.

The excitatory component comprised genes encoding AMPA and NMDA glutamatergic receptor subunits: *GRIA1, GRIA2, GRIA3, GRIA4, GRIN1, GRIN2A, GRIN2B*, and *GRIN2C*. The inhibitory component comprised genes encoding GABA_A_ receptor subunits: *GABRA1, GABRA2, GABRA3, GABRA4, GABRA5, GABRB1, GABRB2, GABRB3, GABRG1, GABRG2*, and *GABRG3*. For each parcel *p*, the transcriptomic E:I ratio was calculated as

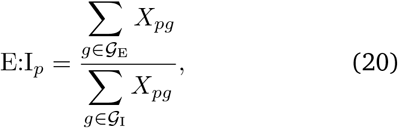

where *X*_*pg*_ denotes the normalized expression of gene *g* in parcel *p*, and *G*_E_ and *G*_I_ denote the excitatory and inhibitory gene sets, respectively. Parcel-wise ratios were min–max scaled to the interval [0, 1] to generate the cortical map. This rescaling does not affect Pearson spatial correlations because it is a positive affine transformation.

Because tissue sampling in the Allen Human Brain Atlas is substantially more complete in the left hemisphere, inferential spatial analyses were restricted to the 500 left-hemisphere Schaefer parcels. The independently estimated bilateral map was retained for cortical visualization and compatibility with the existing whole-cortex processing pipeline; no right-hemisphere values were reflected or otherwise inferred from the left hemisphere.

### Participant-level spatial correlation between E:I balance and turbulence

The bilateral E:I map and each participant’s ComBat-harmonized turbulence map at the largest spatial scale (*λ* = 0.01) were projected from MNI152 space to the fsaverage41k cortical surface using *neuromaps*. Only jointly non-zero left-hemisphere vertices were retained, resulting in 37,254 vertices for the spatial correlations.

For each participant *i*, correspondence with the normative E:I architecture was quantified using the Pearson spatial correlation

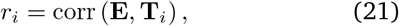

where **E** is the normative left-hemisphere E:I map and T_*i*_ is the participant’s left-hemisphere turbulence map. Correlations were Fisher-transformed before participant-level statistical inference:

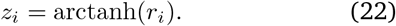

To account for the spatial autocorrelation of cortical maps, we additionally performed a spatial-null sensitivity analysis using Alexander–Bloch rotations. The E:I map was rotated 1,000 times on the fsaverage41k spherical surface. The same set of rotations was used for every participant, while each participant retained an individual null distribution obtained by correlating their turbulence map with each rotated E:I map. Spatially normalized coupling was then calculated as

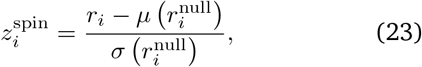

where μ 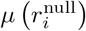 and σ 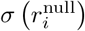 are the mean and standard deviation of participant i’s spatial-null distribution.

### Spatial association between the E:I ratio and turbulence at subject level

Progression of E:I–turbulence spatial alignment across the AD continuum was tested using an ordered-stage regression. Diagnostic stage was coded as HC^*−*^ = 0, HC^+^ = 1, MCI^+^ = 2, and AD^+^ = 3. The model included age, sex, and years of education as covariates:

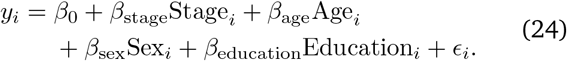

where *y*_*i*_ was the Fisher-transformed spatial correlation at subject-level and the spatially normalized coupling score in each analysis. Confidence intervals were calculated using HC3 heteroscedasticity-robust standard errors. Statistical significance was assessed using 10,000 Freedman–Lane residual permutations.

We performed three categorical contrasts comparing HC− with HC+, MCI+, and AD+ while adjusting for the same covariates. Permutation-derived two-sided *p* values for these contrasts were corrected across the three comparisons using the Benjamini–Hochberg false-discovery-rate procedure.

### Statistical analyses

Demographic and clinical differences across the four amyloid-status groups were evaluated using Kruskal– Wallis tests for continuous variables and Pearson’s *χ*^2^ tests for categorical variables. Significant Kruskal–Wallis effects were followed by Dunn pairwise comparisons with Holm correction. Epsilon squared (*ε*^2^) was used to quantify the effect size of significant main group effects.

Group-level information capability and susceptibility were evaluated from distributions obtained across 100 repeated whole-brain simulation trials for each disease stage. Pairwise group differences were assessed using two-sided permutation tests with 1,000 permutations. Because these measures were derived from group-level simulations rather than individual participants, demographic covariates could not be included. The resulting permutation *p*-values were corrected using the Benjamini–Hochberg false discovery rate (BH-FDR) procedure across the five planned stage comparisons separately for information capability and susceptibility. Group differences in participant-level empirical dynamical measures were assessed using Freedman–Lane residual-permutation tests. For each measure, we fitted the model

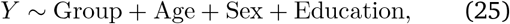

where Group was represented by the four-level amyloid-status classification. Statistical significance was evaluated using 100,000 permutations. Five pairwise comparisons were prespecified: HC^−^ vs. HC^+^, HC^+^ vs. MCI^+^, MCI^+^ vs. AD^+^, HC^−^ vs. MCI^+^, and HC^−^ vs. AD^+^. Pairwise permutation p-values were corrected using BH-FDR across these five contrasts separately within each dynamical measure and spatial scale. Covariate-adjusted Hedges’ *g* was used to quantify pairwise effect sizes.

Participant-level information capability and susceptibility obtained from subject-specific Hopf models were analysed using the same Freedman–Lane framework, with age, sex, and years of education included as nuisance covariates. For each perturbation measure, the main group effect and the five prespecified pairwise contrasts were evaluated using 10,000 permutations. Main group effect *p*-values were corrected across the two perturbation measures, whereas pairwise *p*-values were corrected across the five planned contrasts separately within each measure.

Node-wise metastability was analysed at *λ* = 0.01 using Freedman–Lane residual-permutation testing with age, sex, and education as covariates. The HC^−^ vs. AD^+^ and MCI^+^ vs. AD^+^ contrasts were tested separately at each of the 1,000 Schaefer parcels using 10,000 permutations. A common participant-label permutation was used across nodes within each iteration to preserve the dependence structure among parcels. BH-FDR correction was then applied across the 1,000 nodes separately for each contrast. To characterise the network distribution of the strongest effects, nodes at or below the 30th percentile of the FDR-adjusted permutation *p*-value distribution were selected, retaining ties at the percentile threshold, and assigned to the Yeo seven resting-state networks. Network representation ratios were calculated by dividing the proportion of selected nodes assigned to each network by that network’s proportion of the complete Schaefer atlas.

Cross-sectional associations with MoCA were assessed in separate ordinary least-squares regression models for turbulence, information flow, and information transfer at *λ* = 0.01. Each dynamical predictor was standardised within the analysis sample, such that its regression coefficient represented the expected difference in raw MoCA points per one-standard-deviation increase in the predictor. All models included age, sex, and years of education as covariates and used heteroscedasticity-consistent HC3 standard errors. BH-FDR correction was applied across the three prespecified dynamical predictors. Full-model *R*^2^, adjusted *R*^2^, and the incremental variance explained by the dynamical predictor beyond the demographic covariates (Δ*R*^2^) were calculated.

Because the three empirical dynamical predictors were strongly correlated, their relative contributions to MoCA were additionally examined using an LMG relative-importance decomposition. LMG importance was defined as the average incremental *R*^2^ attributable to each predictor across all six possible predictor-entry orders, while retaining age, sex, and education in every model. Percentile confidence intervals were estimated using 10,000 paired participant-level bootstrap samples. Pairwise bootstrap tests compared the LMG contribution of information flow with those of turbulence and information transfer, with BH-FDR correction across these two comparisons. This analysis was treated as descriptive because the pairwise differences in relative importance did not survive correction.

Associations between the dynamical measures and AT(N) biomarkers were assessed using separate ordinary least-squares regression models for each predictor and outcome. The five prespecified outcomes comprised cortical amyloid burden, bilateral hippocampal grey matter volume, and tau deposition in mesial temporal, metatemporal, and temporoparietal regions. All models included age, sex, and years of education as nuisance covariates and used HC3 heteroscedasticity-consistent standard errors. BH-FDR correction was applied across the five AT(N) outcomes separately for turbulence, information capability, and susceptibility. The regression coefficients reported in the Results correspond to the expected change in the biomarker per raw unit of the respective dynamical predictor. Full-model *R*^2^ and the incremental *R*^2^ attributable to each dynamical predictor beyond the covariates were also calculated. Analyses of amyloid burden and hippocampal volume included 145 participants, whereas regional tau analyses included 134 participants because of differences in tau-PET availability.

## Code and data availability

Data used in the preparation of this article were obtained from the Alzheimer’s Disease Neuroimaging Initiative (ADNI) database (https://adni.loni.usc.edu/). The code developed for this work is publicly available at https://github.com/noechan/hopf_turbu_preprint.

## Data Availability

All data in the present study are available online at https://adni.loni.usc.edu

## ACKNOWLEDGEMENTS

Data collection and sharing by the Alzheimer’s Disease Neuroimaging Initiative (ADNI) are funded by the National Institute on Aging through National Institutes of Health grant U19 AG024904. The grant is administered by the Northern California Institute for Research and Education. Additional historical support for ADNI has been provided by the National Institute of Biomedical Imaging and Bioengineering, the Canadian Institutes of Health Research, and private-sector contributions coordinated through the Foundation for the National Institutes of Health (FNIH).

These private-sector and charitable contributors include AbbVie; Alzheimer’s Association; Alzheimer’s Drug Discovery Foundation; Araclon Biotech; BioClinica, Inc.; Biogen; Bristol-Myers Squibb Company; CereSpir, Inc.; Cogstate; Eisai Inc.; Elan Pharmaceuticals, Inc.; Eli Lilly and Company; EuroImmun; F. Hoffmann-La Roche Ltd and its affiliate Genentech, Inc.; Fujirebio; GE Healthcare; IXICO Ltd.; Janssen Alzheimer Immunotherapy Research Development LLC; Johnson Johnson Pharmaceutical Research Development LLC; Lumosity; Lundbeck; Merck Co., Inc.; Meso Scale Diagnostics, LLC; NeuroRx Research; Neurotrack Technologies; Novartis Pharmaceuticals Corporation; Pfizer Inc.; Piramal Imaging; Servier; Takeda Pharmaceutical Company; and Transition Therapeutics.

This research was undertaken thanks in part to funding from the project eBRAIN-Health - Actionable Multilevel Health Data (id 101058516), funded by the EU Horizon Europe to Noelia Martínez-Molina, Anira Escrichs, Guillermo Montaña-Valverde and Gustavo Deco. David Aquilué-Llorens was supported by a fellowship from the La Caixa” Foundation (ID 100010434), fellowship code B006426. Gustavo Patow was funded by Grant PID2021-122136OB-C22, funded by MI-CIU/AEI/10.13039/501100011033 and by the European Regional Development Fund (ERDF), “A way of making Europe”.

