## Supplementary Material for "Turbulent dynamics and in silico perturbation-based measures are disrupted across biological stages of Alzheimer’s disease"

### SUPPLEMENTARY TABLES

TABLE S1: Pairwise differences in harmonised turbulence across amyloid-status groups after adjustment for age, sex and education.

| Comparison | $\lambda$ | $N_1$ | $N_2$ | $t$ | $p_{\text{perm}}$ | $g_{\text{adj}}$ | $p_{\text{FDR}}$ |
|---|---|---|---|---|---|---|---|
| HC− vs. HC+ | 0.27 | 51 | 37 | -2.15 | 0.03495 | -0.47 | 0.08070 |
| HC− vs. HC+ | 0.24 | 51 | 37 | -0.64 | 0.52575 | -0.14 | 0.82905 |
| HC− vs. HC+ | 0.21 | 51 | 37 | 0.74 | 0.45925 | 0.16 | 0.57406 |
| HC− vs. HC+ | 0.18 | 51 | 37 | 1.68 | 0.09656 | 0.37 | 0.22857 |
| HC− vs. HC+ | 0.15 | 51 | 37 | 2.18 | 0.03151 | 0.48 | 0.15755 |
| HC− vs. HC+ | 0.12 | 51 | 37 | 2.29 | 0.02378 | 0.51 | 0.10440 |
| HC− vs. HC+ | 0.09 | 51 | 37 | 2.12 | 0.03709 | 0.47 | 0.08343 |
| HC− vs. HC+ | 0.06 | 51 | 37 | 1.74 | 0.08606 | 0.38 | 0.14343 |
| HC− vs. HC+ | 0.03 | 51 | 37 | 1.04 | 0.30375 | 0.23 | 0.33740 |
| HC− vs. HC+ | 0.01 | 51 | 37 | 0.49 | 0.62177 | 0.11 | 0.62177 |
| HC+ vs. MCI+ | 0.27 | 37 | 31 | 0.59 | 0.55507 | 0.15 | 0.55507 |
| HC+ vs. MCI+ | 0.24 | 37 | 31 | 0.22 | 0.82905 | 0.05 | 0.82905 |
| HC+ vs. MCI+ | 0.21 | 37 | 31 | -0.07 | 0.94479 | -0.02 | 0.94479 |
| HC+ vs. MCI+ | 0.18 | 37 | 31 | -0.30 | 0.76198 | -0.07 | 0.86670 |
| HC+ vs. MCI+ | 0.15 | 37 | 31 | -0.40 | 0.69364 | -0.10 | 0.69364 |
| HC+ vs. MCI+ | 0.12 | 37 | 31 | -0.34 | 0.73413 | -0.08 | 0.73413 |
| HC+ vs. MCI+ | 0.09 | 37 | 31 | -0.14 | 0.89181 | -0.03 | 0.89269 |
| HC+ vs. MCI+ | 0.06 | 37 | 31 | 0.26 | 0.79575 | 0.06 | 0.79575 |
| HC+ vs. MCI+ | 0.03 | 37 | 31 | 0.97 | 0.33740 | 0.24 | 0.33740 |
| HC+ vs. MCI+ | 0.01 | 37 | 31 | 1.06 | 0.29001 | 0.26 | 0.36251 |
| MCI+ vs. AD+ | 0.27 | 31 | 26 | -2.02 | 0.04842 | -0.57 | 0.08070 |
| MCI+ vs. AD+ | 0.24 | 31 | 26 | -2.14 | 0.03627 | -0.60 | 0.09068 |
| MCI+ vs. AD+ | 0.21 | 31 | 26 | -1.96 | 0.05556 | -0.55 | 0.27780 |
| MCI+ vs. AD+ | 0.18 | 31 | 26 | -1.64 | 0.10601 | -0.46 | 0.22857 |
| MCI+ vs. AD+ | 0.15 | 31 | 26 | -1.25 | 0.21560 | -0.35 | 0.35933 |
| MCI+ vs. AD+ | 0.12 | 31 | 26 | -0.77 | 0.44390 | -0.22 | 0.55488 |
| MCI+ vs. AD+ | 0.09 | 31 | 26 | -0.14 | 0.89269 | -0.04 | 0.89269 |
| MCI+ vs. AD+ | 0.06 | 31 | 26 | 0.85 | 0.39747 | 0.24 | 0.49684 |
| MCI+ vs. AD+ | 0.03 | 31 | 26 | 2.27 | 0.02717 | 0.64 | 0.06792 |
| MCI+ vs. AD+ | 0.01 | 31 | 26 | 2.91 | 0.00512 | 0.82 | **0.01280** |
| HC− vs. MCI+ | 0.27 | 51 | 31 | -1.23 | 0.22342 | -0.28 | 0.27927 |
| HC− vs. MCI+ | 0.24 | 51 | 31 | -0.23 | 0.81728 | -0.05 | 0.82905 |
| HC− vs. MCI+ | 0.21 | 51 | 31 | 0.82 | 0.41424 | 0.19 | 0.57406 |
| HC− vs. MCI+ | 0.18 | 51 | 31 | 1.50 | 0.13714 | 0.35 | 0.22857 |
| HC− vs. MCI+ | 0.15 | 51 | 31 | 1.89 | 0.06344 | 0.44 | 0.15860 |
| HC− vs. MCI+ | 0.12 | 51 | 31 | 2.07 | 0.04176 | 0.48 | 0.10440 |
| HC− vs. MCI+ | 0.09 | 51 | 31 | 2.14 | 0.03475 | 0.49 | 0.08343 |
| HC− vs. MCI+ | 0.06 | 51 | 31 | 2.14 | 0.03519 | 0.49 | 0.08797 |
| HC− vs. MCI+ | 0.03 | 51 | 31 | 1.92 | 0.05701 | 0.44 | 0.09502 |
| HC− vs. MCI+ | 0.01 | 51 | 31 | 1.36 | 0.17854 | 0.31 | 0.29757 |
| HC− vs. AD+ | 0.27 | 51 | 26 | -3.55 | 0.00069 | -0.97 | **0.00345** |
| HC− vs. AD+ | 0.24 | 51 | 26 | -2.50 | 0.01414 | -0.69 | 0.07070 |
| HC− vs. AD+ | 0.21 | 51 | 26 | -1.19 | 0.23842 | -0.33 | 0.57406 |
| HC− vs. AD+ | 0.18 | 51 | 26 | -0.17 | 0.86670 | -0.05 | 0.86670 |
| HC− vs. AD+ | 0.15 | 51 | 26 | 0.62 | 0.53525 | 0.17 | 0.66906 |
| HC− vs. AD+ | 0.12 | 51 | 26 | 1.30 | 0.19784 | 0.36 | 0.32973 |
| HC− vs. AD+ | 0.09 | 51 | 26 | 1.99 | 0.05006 | 0.54 | 0.08343 |
| HC− vs. AD+ | 0.06 | 51 | 26 | 2.84 | 0.00568 | 0.78 | **0.02840** |
| HC− vs. AD+ | 0.03 | 51 | 26 | 3.61 | 0.00064 | 0.99 | **0.00320** |
| HC− vs. AD+ | 0.01 | 51 | 26 | 3.69 | 0.00049 | 1.01 | **0.00245** |

Continued on next page

TABLE S1 – continued from previous page

| Comparison | $\lambda$ | $N_1$ | $N_2$ | $t$ | $p_{\text{perm}}$ | $g_{\text{adj}}$ | $p_{\text{FDR}}$ |
|---|---|---|---|---|---|---|---|

Notes: Results are from Freedman–Lane permutation models adjusted for age, sex and education. $p_{\text{perm}}$ is the two-sided permutation $p$-value. $p_{\text{FDR}}$ is the Benjamini–Hochberg FDR-adjusted $p$-value across the five planned stage comparisons separately within each $\lambda$. The signs of $t$ and adjusted Hedges' $g$ represent the first-named group minus the second-named group; positive values therefore indicate higher turbulence in the first-named group. Bold indicates $p_{\text{FDR}} < 0.05$.

TABLE S2: Pairwise differences in harmonised information flow, information transfer, and information cascade across amyloid-status groups after adjustment for age, sex, and education.

| Comparison | $\lambda$ | $N_1$ | $N_2$ | $t$ | $p_{\text{perm}}$ | $g_{\text{adj}}$ | $p_{\text{FDR}}$ |
|---|---|---|---|---|---|---|---|
| *Information flow* | | | | | | | |
| HC− vs. HC+ | 0.24 | 51 | 37 | 1.57 | 0.12084 | 0.34 | 0.20140 |
| HC− vs. HC+ | 0.21 | 51 | 37 | 1.59 | 0.11443 | 0.35 | 0.19072 |
| HC− vs. HC+ | 0.18 | 51 | 37 | 1.58 | 0.11851 | 0.35 | 0.18643 |
| HC− vs. HC+ | 0.15 | 51 | 37 | 1.40 | 0.16432 | 0.31 | 0.20540 |
| HC− vs. HC+ | 0.12 | 51 | 37 | 1.13 | 0.26203 | 0.25 | 0.26203 |
| HC− vs. HC+ | 0.09 | 51 | 37 | 0.80 | 0.42629 | 0.18 | 0.42629 |
| HC− vs. HC+ | 0.06 | 51 | 37 | 0.41 | 0.68176 | 0.09 | 0.68176 |
| HC− vs. HC+ | 0.03 | 51 | 37 | -0.04 | 0.96638 | -0.01 | 0.96638 |
| HC− vs. HC+ | 0.01 | 51 | 37 | -0.59 | 0.55712 | -0.13 | 0.69640 |
| HC+ vs. MCI+ | 0.24 | 37 | 31 | 1.29 | 0.20288 | 0.32 | 0.25360 |
| HC+ vs. MCI+ | 0.21 | 37 | 31 | 1.38 | 0.17279 | 0.34 | 0.21599 |
| HC+ vs. MCI+ | 0.18 | 37 | 31 | 1.46 | 0.14914 | 0.36 | 0.18643 |
| HC+ vs. MCI+ | 0.15 | 37 | 31 | 1.52 | 0.13283 | 0.37 | 0.20540 |
| HC+ vs. MCI+ | 0.12 | 37 | 31 | 1.53 | 0.13058 | 0.38 | 0.16323 |
| HC+ vs. MCI+ | 0.09 | 37 | 31 | 1.45 | 0.15133 | 0.36 | 0.18916 |
| HC+ vs. MCI+ | 0.06 | 37 | 31 | 1.48 | 0.14364 | 0.36 | 0.17955 |
| HC+ vs. MCI+ | 0.03 | 37 | 31 | 1.92 | 0.05829 | 0.47 | 0.09715 |
| HC+ vs. MCI+ | 0.01 | 37 | 31 | 1.25 | 0.21437 | 0.31 | 0.35728 |
| MCI+ vs. AD+ | 0.24 | 31 | 26 | 0.17 | 0.86388 | 0.05 | 0.86388 |
| MCI+ vs. AD+ | 0.21 | 31 | 26 | 0.37 | 0.71508 | 0.10 | 0.71508 |
| MCI+ vs. AD+ | 0.18 | 31 | 26 | 0.70 | 0.48626 | 0.20 | 0.48626 |
| MCI+ vs. AD+ | 0.15 | 31 | 26 | 1.23 | 0.22408 | 0.35 | 0.22408 |
| MCI+ vs. AD+ | 0.12 | 31 | 26 | 1.95 | 0.05656 | 0.55 | 0.09427 |
| MCI+ vs. AD+ | 0.09 | 31 | 26 | 2.93 | 0.00496 | 0.82 | **0.01240** |
| MCI+ vs. AD+ | 0.06 | 31 | 26 | 3.83 | 0.00023 | 1.08 | **0.00097** |
| MCI+ vs. AD+ | 0.03 | 31 | 26 | 3.90 | 0.00024 | 1.09 | **0.00060** |
| MCI+ vs. AD+ | 0.01 | 31 | 26 | 4.12 | 0.00016 | 1.16 | **0.00080** |
| HC− vs. MCI+ | 0.24 | 51 | 31 | 2.82 | 0.00606 | 0.65 | **0.03030** |
| HC− vs. MCI+ | 0.21 | 51 | 31 | 2.97 | 0.00387 | 0.69 | **0.01935** |
| HC− vs. MCI+ | 0.18 | 51 | 31 | 3.07 | 0.00322 | 0.71 | **0.01610** |
| HC− vs. MCI+ | 0.15 | 51 | 31 | 2.96 | 0.00411 | 0.68 | **0.01027** |
| HC− vs. MCI+ | 0.12 | 51 | 31 | 2.65 | 0.00942 | 0.61 | **0.02355** |
| HC− vs. MCI+ | 0.09 | 51 | 31 | 2.17 | 0.03271 | 0.50 | 0.05452 |
| HC− vs. MCI+ | 0.06 | 51 | 31 | 1.69 | 0.09446 | 0.39 | 0.15743 |
| HC− vs. MCI+ | 0.03 | 51 | 31 | 1.45 | 0.15085 | 0.33 | 0.18856 |
| HC− vs. MCI+ | 0.01 | 51 | 31 | 0.10 | 0.91676 | 0.02 | 0.91676 |
| HC− vs. AD+ | 0.24 | 51 | 26 | 2.19 | 0.03201 | 0.60 | 0.08002 |
| HC− vs. AD+ | 0.21 | 51 | 26 | 2.49 | 0.01445 | 0.68 | **0.03612** |
| HC− vs. AD+ | 0.18 | 51 | 26 | 2.82 | 0.00646 | 0.77 | **0.01615** |
| HC− vs. AD+ | 0.15 | 51 | 26 | 3.11 | 0.00296 | 0.85 | **0.01027** |
| HC− vs. AD+ | 0.12 | 51 | 26 | 3.37 | 0.00131 | 0.92 | **0.00655** |
| HC− vs. AD+ | 0.09 | 51 | 26 | 3.61 | 0.00057 | 0.99 | **0.00285** |
| HC− vs. AD+ | 0.06 | 51 | 26 | 3.74 | 0.00039 | 1.03 | **0.00097** |
| HC− vs. AD+ | 0.03 | 51 | 26 | 3.91 | 0.00020 | 1.07 | **0.00060** |
| HC− vs. AD+ | 0.01 | 51 | 26 | 3.54 | 0.00069 | 0.97 | **0.00172** |
| Information transfer | | | | | | | |
| HC− vs. HC+ | 0.27 | 51 | 37 | 2.02 | 0.04693 | 0.44 | 0.15232 |
| HC− vs. HC+ | 0.24 | 51 | 37 | 1.89 | 0.06164 | 0.42 | 0.15410 |
| HC− vs. HC+ | 0.21 | 51 | 37 | 1.72 | 0.08813 | 0.38 | 0.19422 |
| HC− vs. HC+ | 0.18 | 51 | 37 | 1.51 | 0.13427 | 0.33 | 0.22378 |
| HC− vs. HC+ | 0.15 | 51 | 37 | 1.26 | 0.20879 | 0.28 | 0.34798 |
| HC− vs. HC+ | 0.12 | 51 | 37 | 1.06 | 0.29286 | 0.23 | 0.40339 |
| HC− vs. HC+ | 0.09 | 51 | 37 | 0.86 | 0.38900 | 0.19 | 0.48625 |

Continued on next page

TABLE S2 – continued from previous page

| Comparison | $\lambda$ | $N_1$ | $N_2$ | $t$ | $p_{\mathrm{perm}}$ | $g_{\mathrm{adj}}$ | $p_{\mathrm{FDR}}$ |
|---|---|---|---|---|---|---|---|
| HC− vs. HC+ | 0.06 | 51 | 37 | 0.49 | 0.62376 | 0.11 | 0.62376 |
| HC− vs. HC+ | 0.03 | 51 | 37 | -0.27 | 0.78553 | -0.06 | 0.78553 |
| HC− vs. HC+ | 0.01 | 51 | 37 | -0.93 | 0.35841 | -0.20 | 0.44801 |
| HC+ vs. MCI+ | 0.27 | 37 | 31 | -1.72 | 0.09139 | -0.42 | 0.15232 |
| HC+ vs. MCI+ | 0.24 | 37 | 31 | -1.56 | 0.12359 | -0.38 | 0.19551 |
| HC+ vs. MCI+ | 0.21 | 37 | 31 | -1.35 | 0.18350 | -0.33 | 0.22937 |
| HC+ vs. MCI+ | 0.18 | 37 | 31 | -1.05 | 0.29533 | -0.26 | 0.36916 |
| HC+ vs. MCI+ | 0.15 | 37 | 31 | -0.68 | 0.49823 | -0.17 | 0.49823 |
| HC+ vs. MCI+ | 0.12 | 37 | 31 | -0.28 | 0.78133 | -0.07 | 0.78133 |
| HC+ vs. MCI+ | 0.09 | 37 | 31 | 0.19 | 0.84778 | 0.05 | 0.84778 |
| HC+ vs. MCI+ | 0.06 | 37 | 31 | 0.92 | 0.36063 | 0.23 | 0.45079 |
| HC+ vs. MCI+ | 0.03 | 37 | 31 | 1.46 | 0.14794 | 0.36 | 0.24657 |
| HC+ vs. MCI+ | 0.01 | 37 | 31 | 1.13 | 0.26218 | 0.28 | 0.43697 |
| MCI+ vs. AD+ | 0.27 | 31 | 26 | 1.32 | 0.19199 | 0.37 | 0.23999 |
| MCI+ vs. AD+ | 0.24 | 31 | 26 | 1.44 | 0.15641 | 0.40 | 0.19551 |
| MCI+ vs. AD+ | 0.21 | 31 | 26 | 1.59 | 0.11653 | 0.45 | 0.19422 |
| MCI+ vs. AD+ | 0.18 | 31 | 26 | 1.79 | 0.07965 | 0.50 | 0.19912 |
| MCI+ vs. AD+ | 0.15 | 31 | 26 | 2.02 | 0.04865 | 0.57 | 0.12162 |
| MCI+ vs. AD+ | 0.12 | 31 | 26 | 2.29 | 0.02622 | 0.64 | 0.06555 |
| MCI+ vs. AD+ | 0.09 | 31 | 26 | 2.63 | 0.01149 | 0.74 | **0.02873** |
| MCI+ vs. AD+ | 0.06 | 31 | 26 | 2.99 | 0.00427 | 0.84 | **0.01068** |
| MCI+ vs. AD+ | 0.03 | 31 | 26 | 3.25 | 0.00204 | 0.91 | **0.00510** |
| MCI+ vs. AD+ | 0.01 | 31 | 26 | 3.20 | 0.00242 | 0.90 | **0.01210** |
| HC− vs. MCI+ | 0.27 | 51 | 31 | 0.53 | 0.60168 | 0.12 | 0.60168 |
| HC− vs. MCI+ | 0.24 | 51 | 31 | 0.56 | 0.57379 | 0.13 | 0.57379 |
| HC− vs. MCI+ | 0.21 | 51 | 31 | 0.62 | 0.53617 | 0.14 | 0.53617 |
| HC− vs. MCI+ | 0.18 | 51 | 31 | 0.71 | 0.47928 | 0.16 | 0.47928 |
| HC− vs. MCI+ | 0.15 | 51 | 31 | 0.83 | 0.40459 | 0.19 | 0.49823 |
| HC− vs. MCI+ | 0.12 | 51 | 31 | 0.99 | 0.32271 | 0.23 | 0.40339 |
| HC− vs. MCI+ | 0.09 | 51 | 31 | 1.16 | 0.24753 | 0.27 | 0.41255 |
| HC− vs. MCI+ | 0.06 | 51 | 31 | 1.26 | 0.20962 | 0.29 | 0.34937 |
| HC− vs. MCI+ | 0.03 | 51 | 31 | 0.65 | 0.51586 | 0.15 | 0.64482 |
| HC− vs. MCI+ | 0.01 | 51 | 31 | -0.31 | 0.76369 | -0.07 | 0.76369 |
| HC− vs. AD+ | 0.27 | 51 | 26 | 1.89 | 0.06354 | 0.52 | 0.15232 |
| HC− vs. AD+ | 0.24 | 51 | 26 | 2.03 | 0.04558 | 0.56 | 0.15410 |
| HC− vs. AD+ | 0.21 | 51 | 26 | 2.21 | 0.02954 | 0.61 | 0.14770 |
| HC− vs. AD+ | 0.18 | 51 | 26 | 2.44 | 0.01688 | 0.67 | 0.08440 |
| HC− vs. AD+ | 0.15 | 51 | 26 | 2.68 | 0.00915 | 0.74 | **0.04575** |
| HC− vs. AD+ | 0.12 | 51 | 26 | 2.96 | 0.00402 | 0.81 | **0.02010** |
| HC− vs. AD+ | 0.09 | 51 | 26 | 3.31 | 0.00153 | 0.91 | **0.00765** |
| HC− vs. AD+ | 0.06 | 51 | 26 | 3.63 | 0.00063 | 1.00 | **0.00315** |
| HC− vs. AD+ | 0.03 | 51 | 26 | 3.44 | 0.00108 | 0.94 | **0.00510** |
| HC− vs. AD+ | 0.01 | 51 | 26 | 2.82 | 0.00606 | 0.77 | **0.01515** |
| *Information cascade* | | | | | | | |
| HC− vs. HC+ | – | 51 | 37 | 0.08 | 0.93773 | 0.02 | 0.93773 |
| HC+ vs. MCI+ | – | 37 | 31 | 1.62 | 0.11042 | 0.40 | 0.18403 |
| MCI+ vs. AD+ | – | 31 | 26 | 4.10 | 0.00014 | 1.15 | **0.00035** |
| HC− vs. MCI+ | – | 51 | 31 | 1.27 | 0.20598 | 0.29 | 0.25748 |
| HC− vs. AD+ | – | 51 | 26 | 3.98 | 0.00014 | 1.09 | **0.00035** |

Notes: Results are from Freedman–Lane permutation models with 100,000 permutations, adjusted for age, sex and education. Information transfer is analysed after transforming each harmonised value as $1 - x$. $p_{\mathrm{perm}}$ is the two-sided permutation $p$-value. $p_{\mathrm{FDR}}$ is the Benjamini–Hochberg FDR-adjusted $p$-value across the five planned stage comparisons separately within each spatial scale and measure. Information cascade is a scalar summary, so $\lambda$ is not applicable. The signs of $t$ and adjusted Hedges' $g$ represent the first-named group minus the second-named group. Bold indicates $p_{\mathrm{FDR}} < 0.05$.

TABLE S3. Classification performance using linear and polynomial-kernel support vector machine (SVM) classifiers for turbulence-derived features and combined turbulence- and perturbation-derived features across disease-stage contrasts. Values are reported as mean ± standard deviation across 40 resampling iterations. The number of selected features is the optimal number selected within each iteration, summarized across iterations; performance metrics are expressed as percentages.

| **Classifier** | **Feature Set** | **N Selected Features** | **ROC-AUC (Test)** | **Balanced Accuracy (Test)** | **Sensitivity (Test)** | **Specificity (Test)** |
|---|---|---|---|---|---|---|
| | | **HC (A$\beta^-$) vs. AD (A$\beta^+$)** | | | | |
| Linear SVM | Turbulence | $8.50 \pm 2.90$ | $82.80 \pm 9.47$ | $73.06 \pm 8.19$ | $64.14 \pm 18.73$ | $81.98 \pm 11.85$ |
| Linear SVM | Turbulence + Perturbation | $8.00 \pm 2.46$ | $85.43 \pm 5.65$ | $69.29 \pm 9.32$ | $56.20 \pm 15.46$ | $82.37 \pm 17.77$ |
| Polynomial SVM | Turbulence | $8.43 \pm 2.84$ | $74.65 \pm 13.44$ | $66.35 \pm 14.67$ | $59.62 \pm 25.42$ | $73.09 \pm 11.58$ |
| Polynomial SVM | Turbulence + Perturbation | $6.38 \pm 2.41$ | $85.61 \pm 5.89$ | $77.95 \pm 5.66$ | $74.29 \pm 13.08$ | $81.60 \pm 9.12$ |
| | | **HC (A$\beta^-$) vs. MCI (A$\beta^+$)** | | | | |
| Linear SVM | Turbulence | $10.98 \pm 2.84$ | $70.40 \pm 8.90$ | $65.31 \pm 8.64$ | $66.53 \pm 18.92$ | $64.10 \pm 19.54$ |
| Linear SVM | Turbulence + Perturbation | $11.73 \pm 2.80$ | $72.99 \pm 12.45$ | $64.18 \pm 9.80$ | $57.06 \pm 20.95$ | $71.29 \pm 14.60$ |
| Polynomial SVM | Turbulence | $7.73 \pm 2.66$ | $61.63 \pm 10.82$ | $60.51 \pm 4.13$ | $57.79 \pm 4.89$ | $63.23 \pm 8.33$ |
| Polynomial SVM | Turbulence + Perturbation | $10.13 \pm 4.19$ | $66.35 \pm 12.30$ | $59.23 \pm 9.19$ | $54.33 \pm 26.31$ | $64.13 \pm 15.04$ |
| | | **MCI (A$\beta^+$) vs. AD (A$\beta^+$)** | | | | |
| Linear SVM | Turbulence | $9.65 \pm 2.28$ | $77.87 \pm 7.62$ | $66.43 \pm 15.19$ | $44.56 \pm 29.73$ | $88.31 \pm 10.66$ |
| Linear SVM | Turbulence + Perturbation | $8.53 \pm 3.74$ | $79.93 \pm 12.15$ | $70.23 \pm 10.90$ | $63.14 \pm 21.57$ | $77.32 \pm 12.46$ |
| Polynomial SVM | Turbulence | $7.80 \pm 2.77$ | $63.86 \pm 18.65$ | $61.61 \pm 9.44$ | $52.42 \pm 28.52$ | $70.80 \pm 15.76$ |
| Polynomial SVM | Turbulence + Perturbation | $8.10 \pm 2.62$ | $67.35 \pm 25.53$ | $65.98 \pm 11.57$ | $46.06 \pm 32.23$ | $85.90 \pm 15.13$ |
| | | **HC (A$\beta^-$) vs. HC (A$\beta^+$)** | | | | |
| Linear SVM | Turbulence | $10.53 \pm 3.13$ | $62.78 \pm 12.47$ | $58.90 \pm 7.06$ | $58.21 \pm 22.75$ | $59.59 \pm 24.27$ |
| Linear SVM | Turbulence + Perturbation | $9.88 \pm 3.37$ | $52.87 \pm 19.50$ | $58.26 \pm 12.53$ | $51.43 \pm 25.25$ | $65.09 \pm 21.65$ |
| Polynomial SVM | Turbulence | $13.85 \pm 2.44$ | $65.67 \pm 8.13$ | $55.46 \pm 6.21$ | $30.33 \pm 15.96$ | $80.60 \pm 15.04$ |
| Polynomial SVM | Turbulence + Perturbation | $9.60 \pm 3.22$ | $57.18 \pm 17.21$ | $57.47 \pm 9.96$ | $48.91 \pm 31.25$ | $66.03 \pm 23.38$ |

TABLE S4. Spatial correlations between cortical AD-related gene expression and group differences in turbulent dynamics at $\lambda = 0.01$. Cortical gene-expression maps were correlated with the absolute group-mean turbulence-difference maps for the $HC^-$ vs. $AD^+$ and $MCI^+$ vs. $AD^+$ contrasts. Pearson correlation coefficients ($r$) and spatial permutation $p$-values ($p_{spin}$) were estimated using 1,000 Alexander–Bloch rotations on the fsaverage-41k surface. Bold values indicate nominal $p_{spin} < 0.05$. The $p_{spin}$ values were not adjusted for multiple comparisons across genes.

| | **HC⁻ vs. AD⁺** | | **MCI⁺ vs. AD⁺** | |
|---|---|---|---|---|
| **Gene** | $r$ | $p_{spin}$ | $r$ | $p_{spin}$ |
| APP | 0.313 | < 0.001 | 0.261 | < 0.001 |
| ADAM10 | 0.305 | < 0.001 | 0.032 | 0.755 |
| BACE1 | 0.225 | **0.013** | 0.322 | < 0.001 |
| PSEN1 | 0.289 | < 0.001 | 0.047 | 0.391 |
| PSEN2 | 0.223 | **0.036** | 0.413 | < 0.001 |
| APOE | 0.303 | **0.010** | 0.395 | **0.012** |
| CLU | 0.183 | 0.109 | 0.500 | < 0.001 |
| SORL1 | 0.147 | 0.303 | 0.023 | 0.927 |
| ABCA7 | 0.157 | **0.040** | -0.071 | 0.532 |
| BIN1 | 0.298 | < 0.001 | 0.185 | **0.002** |
| CD2AP | 0.210 | < 0.001 | -0.054 | 0.484 |
| CD33 | 0.272 | **0.008** | 0.418 | < 0.001 |
| PTK2B | 0.348 | < 0.001 | 0.126 | 0.170 |
| PICALM | 0.290 | < 0.001 | 0.048 | 0.431 |
| RIN3 | 0.233 | < 0.001 | 0.287 | < 0.001 |

### SUPPLEMENTARY FIGURES

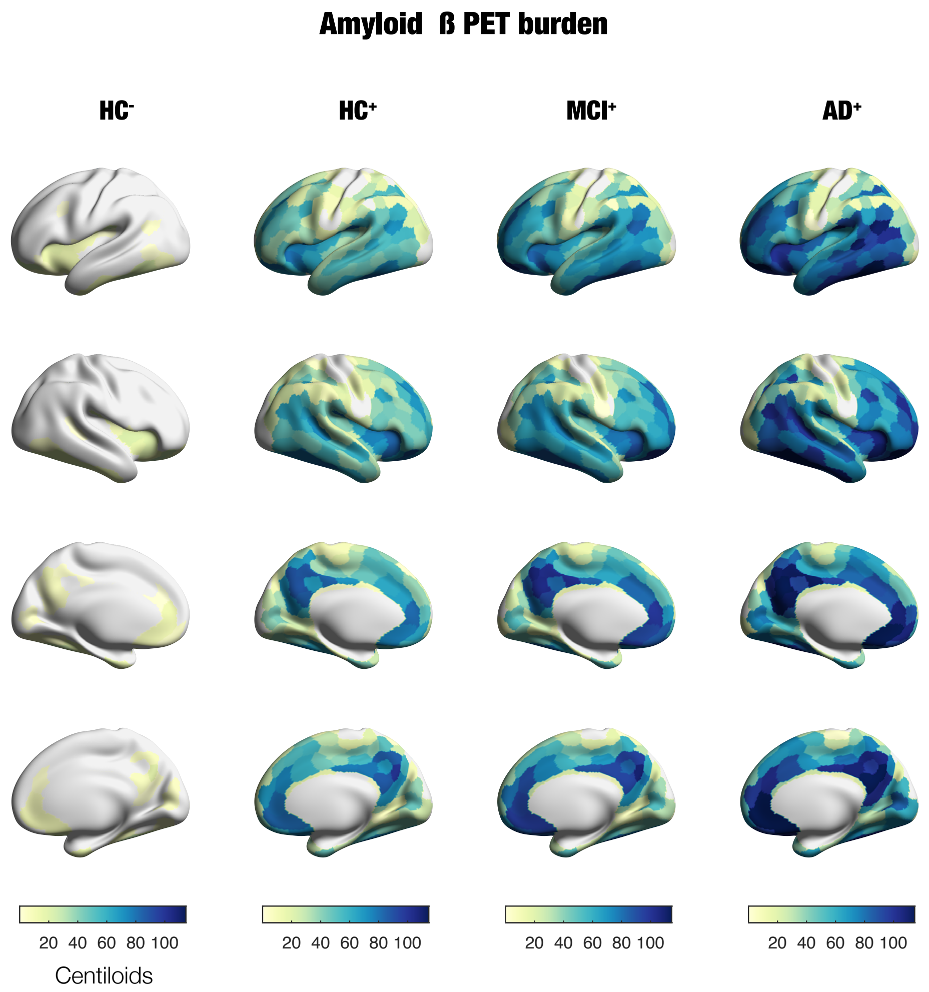


FIG. S1. **Cortical amyloid-$\beta$ PET burden across diagnostic groups.** Brain renderings show the mean regional amyloid-$\beta$ burden for amyloid-negative healthy controls (HC$^-$), amyloid-positive healthy controls (HC$^+$), amyloid-positive participants with mild cognitive impairment (MCI$^+$), and amyloid-positive participants with Alzheimer's disease (AD$^+$). Amyloid burden is expressed on the Centiloid scale. Lateral and medial views of both hemispheres are displayed for each group.

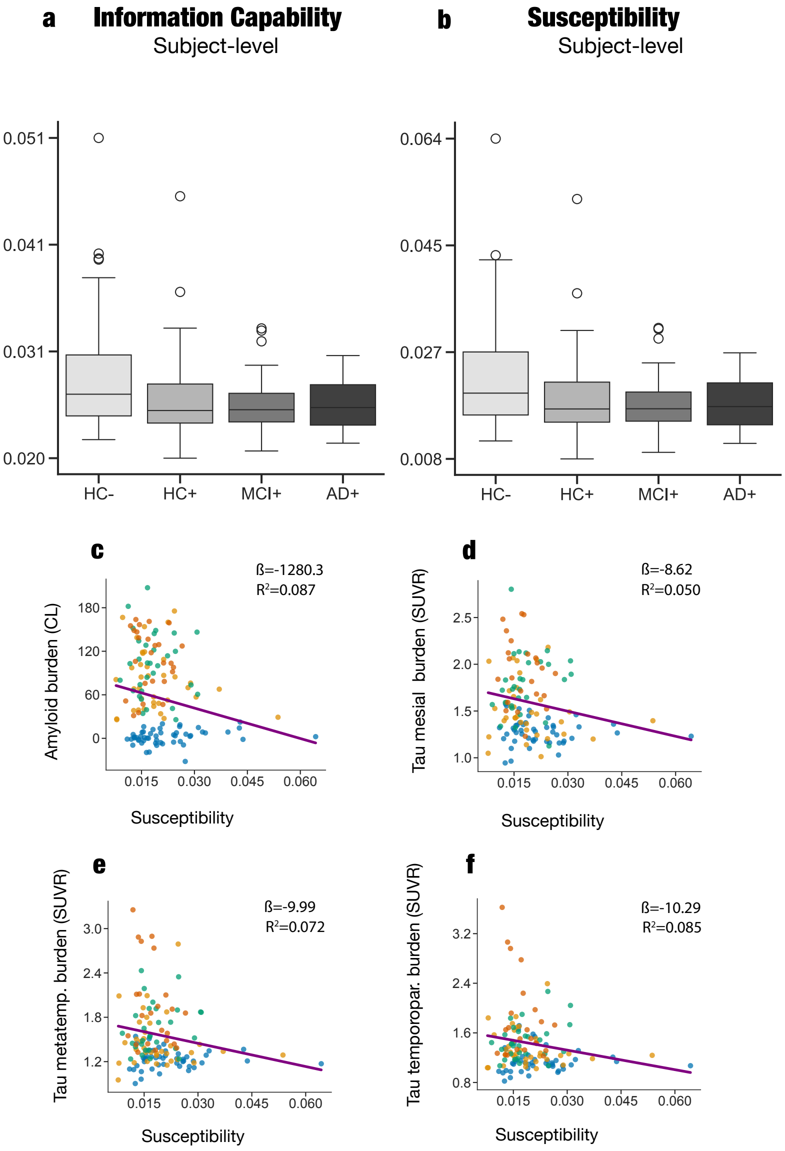


FIG. S2. **Participant-level perturbation measures and their associations with amyloid and tau burden.** (a,b) Participant-level information capability (a) and susceptibility (b), estimated from subject-specific whole-brain Hopf models, across $HC^-$, $HC^+$, $MCI^+$, and $AD^+$ groups. Boxes indicate the median and interquartile range, whiskers extend to 1.5 times the interquartile range, and open circles denote observations beyond the whiskers. (c–f) Associations between susceptibility and cortical amyloid-$\beta$ burden in Centiloid units (c), mesial temporal tau burden (d), metatemporal tau burden (e), and temporoparietal tau burden (f). Tau burden is expressed as standardised uptake value ratio (SUVR). Points represent individual participants and colours indicate diagnostic group. Solid lines show the fitted association. Regression coefficients ($\beta$) and full-model $R^2$ values are reported within each panel. Statistical models were adjusted for age, sex, and years of education.

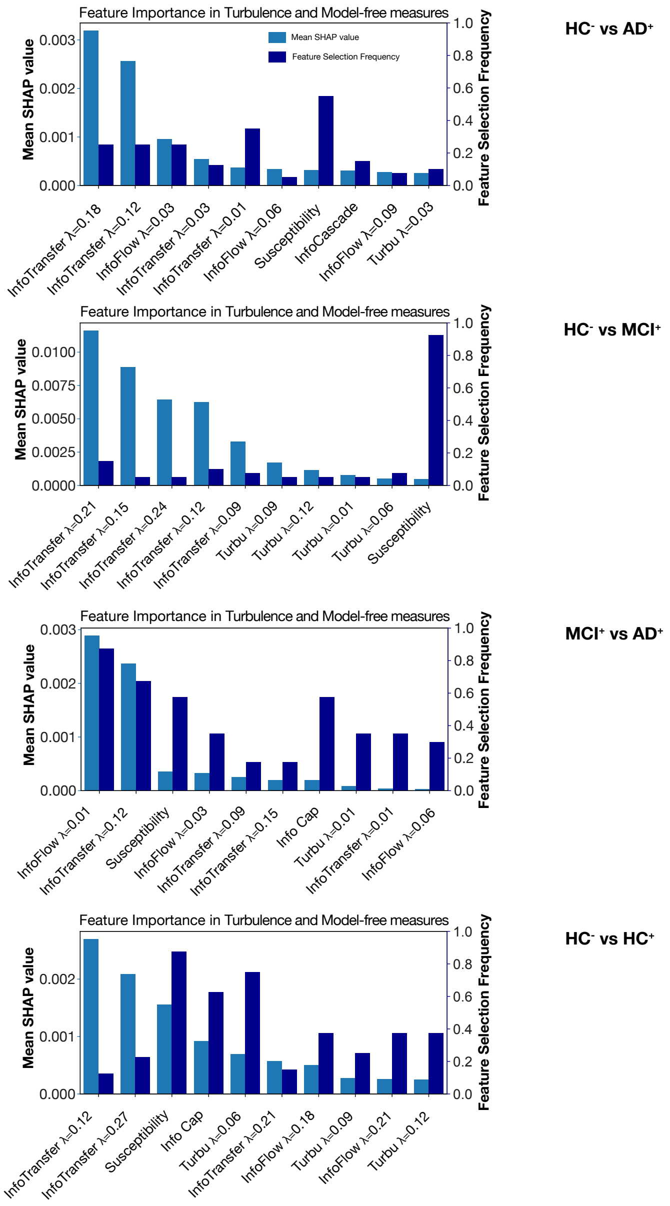


FIG. S3. **Feature importance in classifiers combining empirical turbulent-dynamics and model-based perturbation measures.** Feature importance is shown for the $HC^-$ vs. $AD^+$, $HC^-$ vs. $MCI^+$, $MCI^+$ vs. $AD^+$, and $HC^-$ vs. $HC^+$ classifications, from top to bottom. Light-blue bars show the mean absolute SHapley Additive exPlanations (SHAP) value for each feature in the held-out test samples, aggregated across 40 repeated train-test partitions. Dark-blue bars show the proportion of repetitions in which each feature was selected. Empirical features comprise information flow, information transfer, information cascade, and turbulence at the indicated spatial scales ($\lambda$); information capability and susceptibility were derived from subject-specific whole-brain Hopf models. Features are ordered by mean absolute SHAP value within each contrast.

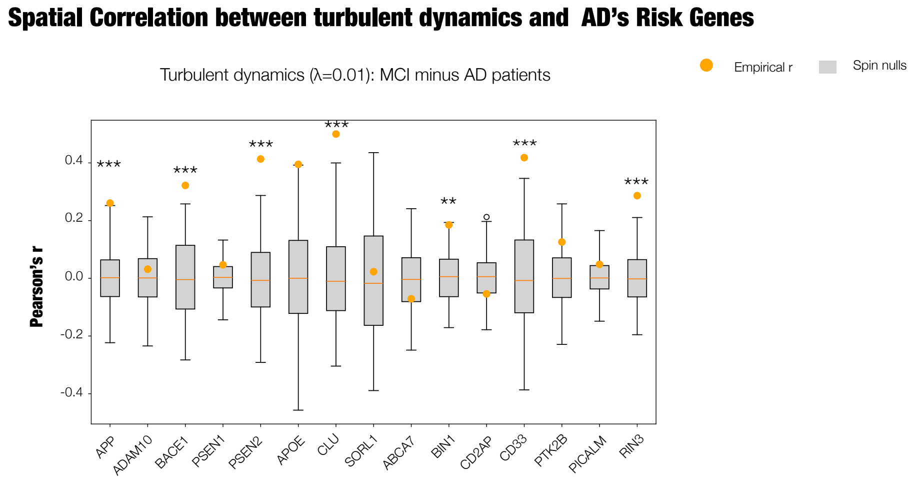


FIG. S4. **Spatial correlations between altered turbulent dynamics and Alzheimer's disease-risk gene-expression patterns in MCI$^+$ versus AD$^+$.** Orange points show the empirical Pearson correlation coefficient ($r$) between the absolute MCI$^+$–AD$^+$ group-mean difference in turbulent dynamics at $\lambda = 0.01$ and the cortical expression map of each Alzheimer's disease-risk gene. Grey boxplots show the corresponding null distributions generated using 1,000 Alexander–Bloch spin rotations of the turbulence-difference map on the fsaverage-41k cortical surface. Asterisks indicate spatial-permutation significance ($^{**}p_{\rm spin} < 0.01$, $^{***}p_{\rm spin} < 0.001$).

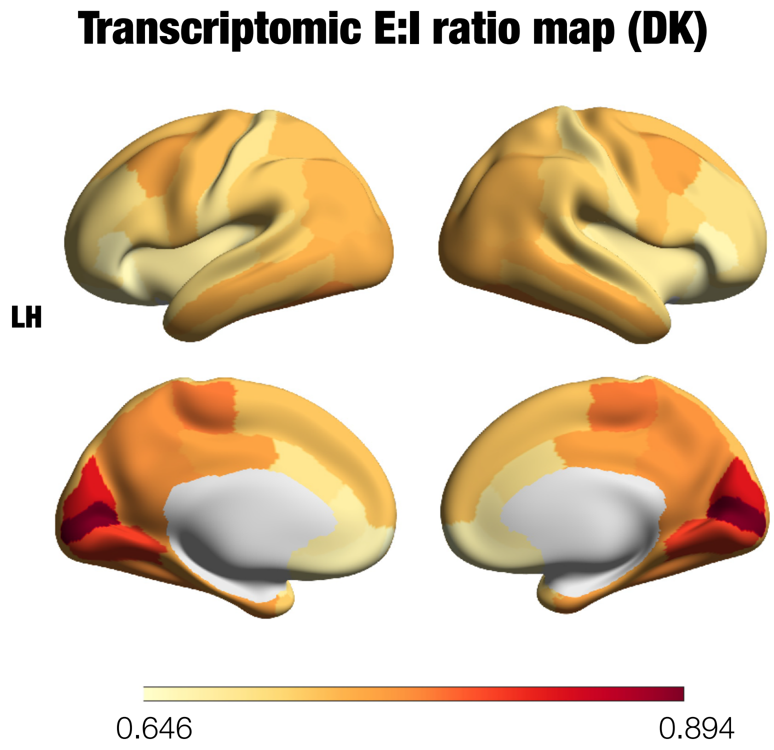


FIG. S5. **Maps of E:I ratio with the Desikan-Killiany parcellation**. E:I is the ratio of excitatory to inhibitory receptor gene expression. See Methods for further details.
